# Stillbirths: contribution of preterm birth and size-for-gestational age for 119.6 million total births from nationwide records in 12 countries, 2000 to 2020

**DOI:** 10.1101/2023.04.14.23288565

**Authors:** Y. B. Okwaraji, L. Suárez-Idueta, E.O. Ohuma, E. Bradley, J. Yargawa, V. Pingray, G. Cormick, A. Gordon, V. Flenady, E. Horváth-Puhó, H.T. Sørensen, L. Sakeus, L. Abuladze, M. Heidarzadeh, N. Khalili, K. A. Yunis, A. Al Bizri, SD. Karalasingam, J. Ravichandran, A. Barranco, Aimée E. van Dijk, L. Broeders, F.F. Alyafei, M. AlQubaisi, N. Razaz, J. Söderling, L. K. Smith, R. J. Matthews, R. Wood, K. Monteath, I. Pereyra, G. Pravia, S. Lisonkova, Q. Wen, J. E. Lawn, H. Blencowe, the National Vulnerable Newborn Collaborative Group for stillbirths

## Abstract

**Objective:** To examine the contribution of preterm birth and size-for-gestational age in stillbirths using six ‘newborn types’.

**Design:** Population-based multi-country analyses.

**Setting:** Births collected through routine data systems in 12 countries.

**Sample:** 119,644,788 total births from 22^+0^ to 44^+6^ weeks gestation identified from 2000 to 2020.

**Methods:** We included 605,557 stillbirths from 22^+0^ weeks gestation from 12 countries. We classified all births, including stillbirths, by six ‘newborn types’ based on gestational age information (preterm, PT, <37^+0^ weeks vs term, T, ≥37^+0^ weeks) and size-for-gestational age defined as small (SGA, <10^th^ centile), appropriate (AGA, 10^th^-90^th^ centiles), or large (LGA, >90^th^ centile) for gestational age, according to the international newborn size for gestational age and sex INTERGROWTH-21^st^ standards.

**Main Outcome Measures:** Distribution of stillbirths, stillbirth rates and rate ratios according to six newborn types.

**Results:** 605,557 (0.50%) of the 119,644,788 total births resulted in stillbirth after 22^+0^ weeks. Most stillbirths (74.3%) were preterm. Around 21.0% were SGA types (PT+SGA (16.0%), T+SGA (5.0%)) and 14.3% were LGA types (PT+LGA (10.1%), T+LGA (4.2%)). The median rate ratio (RR) for stillbirth was highest in PT+SGA babies (RR=78.8, interquartile range (IQR), 68.2, 111.5) followed by PT+AGA (RR=24.5, IQR, 19.3, 29.4), PT+LGA (RR=23.0, IQR,13.7, 29.0) and T+SGA (RR=5.5, IQR, 5.0, 6.0) compared with T+AGA. Stillbirth rate ratios were similar for T+LGA vs T+AGA (RR=0.7, IQR, 0.7, 1.1). At the population level, 21.5% of stillbirths were attributable to small-for-gestational-age.

**Conclusions:** In these high-quality data from high/middle income countries, almost three quarters of stillbirths were born preterm and a fifth were small-for-gestational age, with the highest stillbirth rates associated with the coexistence of preterm and SGA. Further analyses are needed to better understand patterns of gestation-specific risk in these populations, and also patterns in lower-income contexts, especially those with higher rates of intrapartum stillbirth and SGA.

**Funding:** The Children’s Investment Fund Foundation, 1803-02535

**KEY FINDINGS:** *WHAT WAS KNOWN?:* Stillbirth (pregnancy loss after 22^+0^ weeks) is a devastating outcome. Global estimates indicating 1.9 million late gestation stillbirths (≥28^+0^ weeks) worldwide in 2021 underestimate the overall burden. Many of the pathways to stillbirth result in fetal death before term (preterm stillbirth, <37^+0^ weeks of gestational age). In addition, babies with fetal growth restriction (frequently assessed using the proxy small for gestational age (SGA, <10^th^ centile)) are at higher risk of stillbirth than their appropriately grown peers. Stillbirths are therefore more likely to be low birthweight (LBW, <2,500g). Being large for gestational age (LGA, >90^th^ centile) at term may also be associated with increased risk of stillbirth.

*WHAT WAS DONE THAT IS NEW?:* Combining information on gestational age (preterm (PT), or term (T)) and attained size for-gestational-age (small-for-gestational-age (SGA), appropriate-for-gestational age (AGA), large-for-gestational age (LGA)) we defined six ‘newborn types’: four small (PT+SGA, PT+AGA, PT+LGA, T+SGA), one large (T+LGA), and one reference (T+AGA). We compiled livebirth and stillbirth data from 15 high- and middle-income countries as part of the Vulnerable Newborn Collaboration. A total of 119,039,231 livebirths and 605,557 stillbirths ≥22^+0^ weeks from 12 countries between 2000 and 2020 met the inclusion criteria. We examined the distribution of stillbirths by these ‘newborn types’, and calculated type-specific stillbirth rates and rate ratios.

*WHAT WAS FOUND?:* Most stillbirths (74.3%) were preterm, compared to fewer than 1-in-10 (9.0%) livebirths. A fifth (21.0%) of stillbirths were SGA compared to 1-in-20 (5.4%) livebirths. Preterm SGA had 78.8 times higher stillbirth rates compared to term AGA (Rate ratio (RR)=78.8, interquartile range (IQR), 68.2,111.5). Overall, preterm types had a 25 times higher stillbirth rate than term types (RR=25.0, IQR,20.1, 29.5). At the population level, over a fifth of stillbirths (21.5%) were attributable to being SGA, indicating a substantial impact of growth restriction on stillbirth in these settings. 14.3% of stillbirths and 17.5% of livebirths were LGA. There was no evidence of increased stillbirth rates for LGA types. The distribution of these ‘newborn types’ are similar amongst stillbirths and neonatal deaths.

*WHAT NEXT?:* Categorisation of all births, including stillbirths, into these ‘newborn types’ was analytically possible using routinely collected data in these 12 upper-middle- or high-income contexts and led to programmatic relevant findings. However, as the majority (98%) of the world’s stillbirths are in low-and middle-income countries, more data are needed to improve understanding of patterns in stillbirths in a wider range of contexts, especially in settings with higher rates of intrapartum stillbirth and those with very high SGA rates such as South Asia. Further analyses, including assessing gestational age-specific risk, could provide more information on pathways to stillbirth and enable targeted interventions to underlying causes such as infection and obstetric complications. When analysing these vulnerability pathways, omitting stillbirths neglects an important part of the burden and its effects on families and society.

## INTRODUCTION

The World Health Organization (WHO) defines stillbirth as the loss of a baby during pregnancy at or after 22^+0^ weeks of gestation, or if gestational age is not available, weighing 500g or more.^1^ Global estimates are only available for late gestation stillbirths. These estimate that 1.9 million babies were stillborn after 28^+0^ weeks gestation in 2021.^2^ Stillbirth is associated with large emotional toll on affected women, families, health workers and society, representing a substantial loss of human capital.^3^ Importantly, most of these deaths are preventable through improved access to high-quality antenatal and intrapartum care.^4,5^

The Every Newborn Action Plan set a target of 12 or fewer late gestation stillbirths per 1000 total births by 2030.^6,7^ According to the latest estimates, if current trends persist, 56 countries will not meet this stillbirth rate target by 2030.^2,8^ The countries needing most acceleration to meet these targets are in sub-Saharan Africa and South Asia, where stillbirth rates are highest, yet data availability is lowest. Further epidemiological data are needed to understand drivers of stillbirth to inform investments for programmatic action towards ending these frequently preventable deaths.^6^ Data on stillbirths are now available from 173 countries (with data from 138 countries meeting quality inclusion criteria for UN estimates). Many middle- and higher-income countries have individual-level data records which can enable more detailed assessments, which could lead to insights in patterns of stillbirth to inform interventions.

Stillborn babies are more likely to be growth restricted (assessed at birth using the proxy of small for gestational age (SGA, <10^th^ centile)) or preterm (<37^+0^ weeks of gestational age) and therefore more likely to be low birthweight (LBW, <2500g) than liveborn peers.^9,10^ Previous studies have shown that babies compromised through poor fetal growth are at higher risk of stillbirth – both prior to the start of labour (antepartum stillbirth) and during labour (intrapartum stillbirth).^11,12^

LBW has traditionally been used as the main marker of vulnerability. Recent work recognising the two underlying pathways to LBW – short gestation and fetal growth restriction – has proposed the concept of vulnerable ‘newborn types’, with an initial focus primarily on livebirths.^13,14^ No studies to date have sought to categorise stillbirths using these types.

Ashorn et al called for a better description of the prevalence and mortality risk of ‘newborn types’ based on length of gestation and size for gestational age at birth to delineate vulnerability.^13^ These ‘newborn types’ could also assist in the identification of babies at the highest risk of complications, to help better understand biological mechanisms, to inform more targeted and innovative interventions, and to accelerate progress towards global LBW and neonatal mortality reduction targets. Accompanying papers in this supplement have described the prevalence and mortality risk by ‘newborn type’ amongst livebirths.^15,16^ These have demonstrated the relationship between newborn type and neonatal mortality risk with the greatest risk for preterm ‘newborn types’, especially with co-existence of preterm and SGA.

This paper aims to assess the use of this classification to provide a more granular description of stillbirths. In this study, we examined the distribution of stillbirths by these ‘newborn types’.

## METHODS

### Data source

A detailed description of how data were collated is published in detail elsewhere.^14,16^ In brief, 14 of the 23 countries participating in the Vulnerable Newborn Measurement collaboration provided information on stillbirths and were considered in these analyses. Data from the 14 countries were compiled for all births (livebirths and stillbirths) from 2000 to 2020, including 147 country-years. Each country team analysed their datasets using standardised codes in statistical programs Stata, R or SAS using programming developed centrally by the London School of Hygiene & Tropical Medicine (LSHTM), with summary tables shared online through a secured data hub. In accordance with the International Classification of Diseases, stillbirths were defined as fetal deaths at ≥22^+0^ weeks of gestation (Table S1a).^1^ Sensitivity analyses were undertaken to include only late gestation stillbirths at ≥28^+0^ weeks of gestation.

Individual birth records missing birthweight, gestational age and/or sex were excluded as it was not possible to assess size-for-gestational age (Figure 1a). Birth records with gestational age <22^+0^ weeks or >44^+6^ weeks or implausible combinations of birthweight and gestational age (defined as birthweight ±5 standard deviations from the mean birthweight for gestational age) were also excluded.

**Figure 1.**
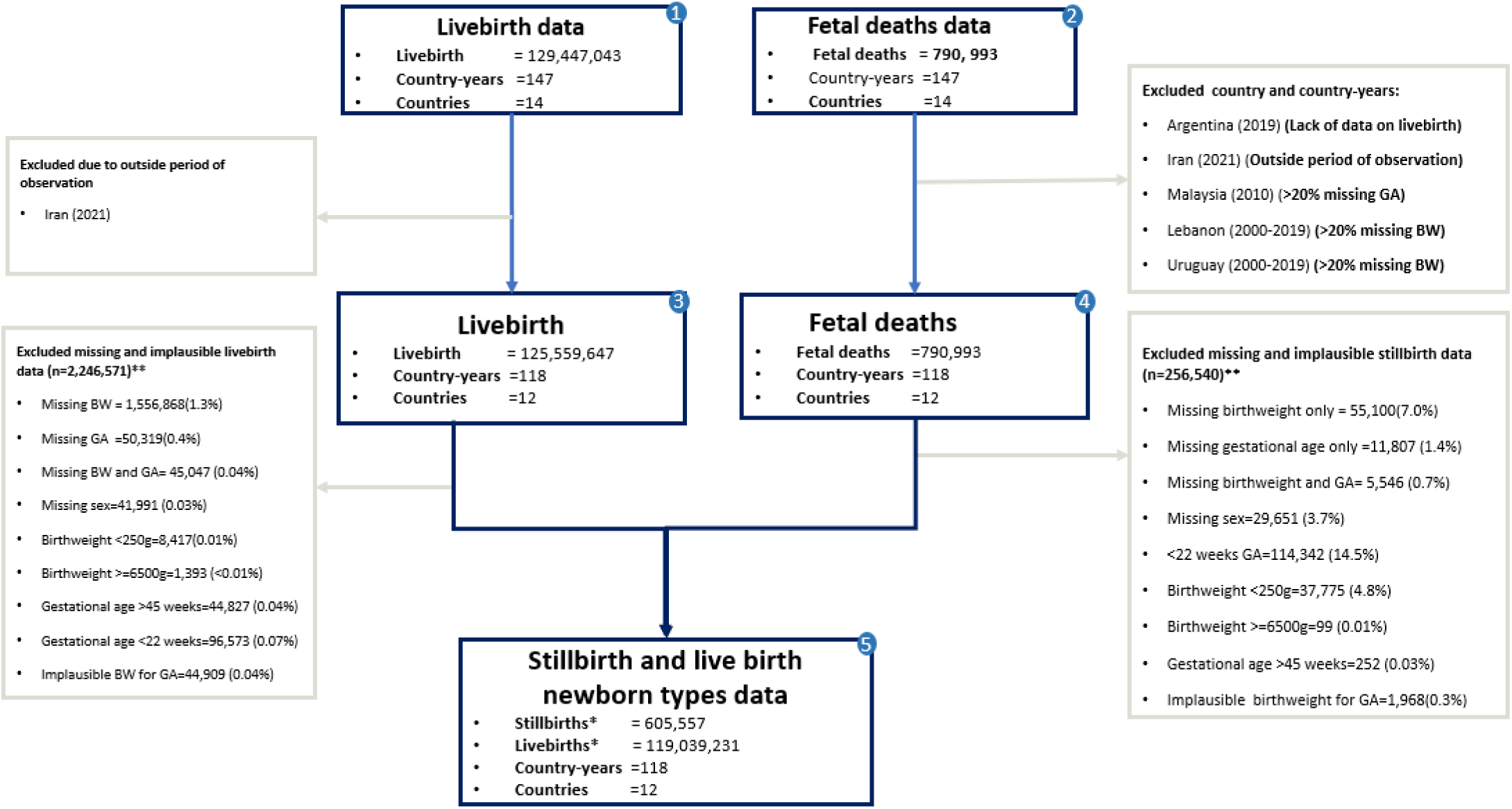
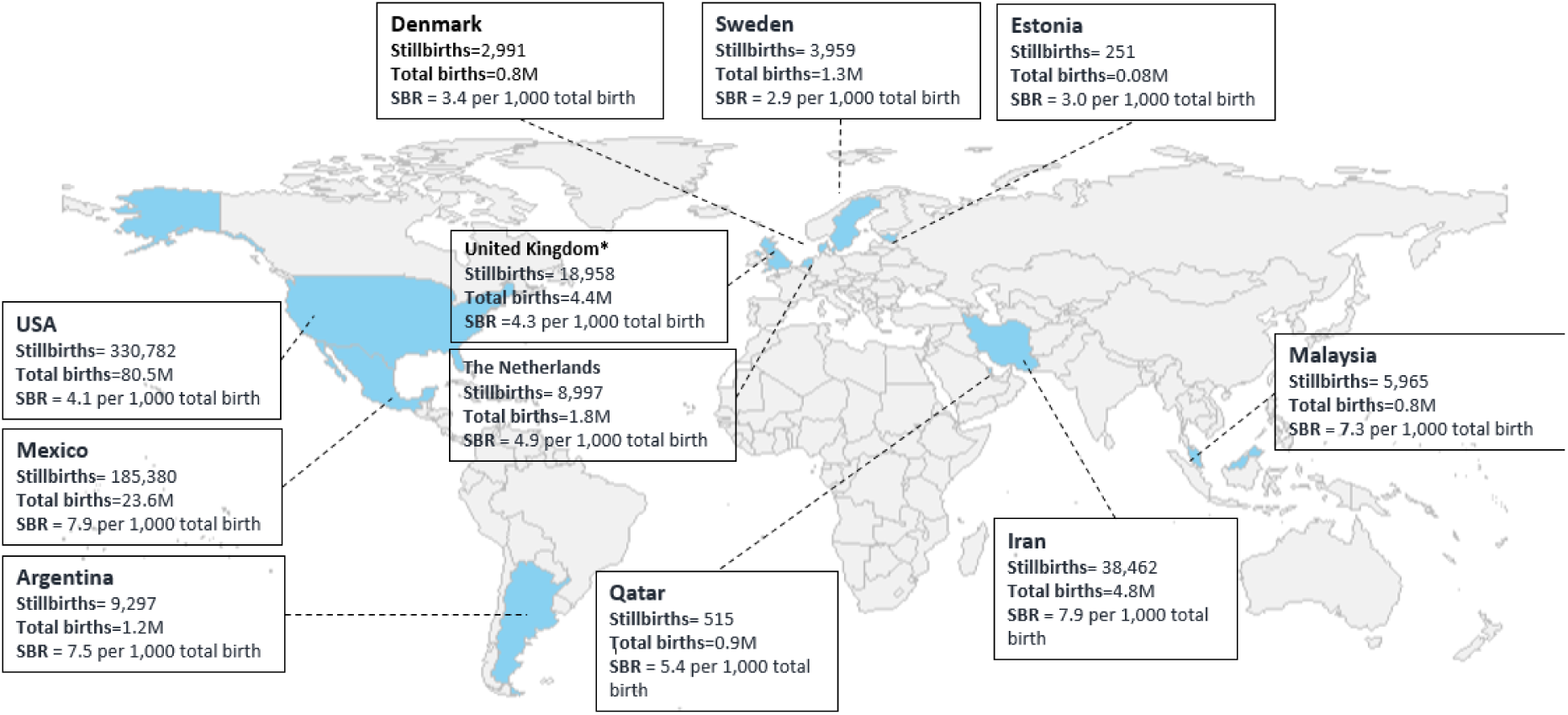
Input data for stillbirth analyses, 2000 to 2020. **Figure 1a. Flowchart of data inclusions and exclusions** *For the sensitivity analysis: 218,351 stillbirths and 590,501 livebirths with GA 22 - 27 weeks were excluded. Total number of births 28 or more weeks: *387,206* late gestation stillbirths, *118,448,730* livebirths * \**Due to overlaps of missing and implausible data, the total excluded values do not odd up to the difference between box 3 and box 4 and between box 3 and box 5*. **Figure 1b. Number of stillbirths (605,557) and total births (119.6 million) by country** *Map legend shows the distribution of 119.6 million total births (119,039,231 livebirths and 605,557 stillbirths at ≥22*^*+0*^ *weeks) with information to classify by ‘newborn types’ included in this study*.

Data quality assessments were performed by estimating the level of missingness of core variables and of implausible values by each country-year (Table S1b). We evaluated the plausibility of the stillbirth dataset by comparing the absolute differences between the calculated late gestation stillbirth rate (SBR) (≥28^+0^ weeks) in our data and the nationally reported SBR for late gestation stillbirth rates (Table S1c).^8^ We excluded country-years with >20% missing birthweight or gestational age data.

Findings are reported in accordance with the Reporting guidelines of studies Conducted using Observational Routinely collected Data, the RECORD statement (Table S2). Ethics approval for all participants is presented as supporting information (Table S3).

### Construction of ‘newborn types’ as exposure indicators

Consistent with the approach previously taken for livebirths,^16,17^ each birth was categorised into six mutually exclusive ‘newborn types’. First, we categorised every birth record as preterm (<37^+0^ weeks (PT)) or term (≥37^+0^ weeks (T)). Next, we classified births by size-for-gestational age defined as small (SGA, <10^th^ centile), appropriate (AGA, 10^th^-90^th^ centiles), or large (LGA, >90^th^ centile) for gestational age using a modified version of the INTERGROWTH-21^st^ international newborn size for gestational age and sex standards extended to include all births from 22^+0^ to 44^+6^ weeks.^18^ We created a set of a six ‘newborn types’ based on the combination of PT or T and size-for-gestational age: four small (PT+SGA, PT+AGA, PT+LGA, T+SGA), one large (T+LGA), and one reference (T+AGA).

### Data Statistical analysis

Among the included records measures were calculated and summarised with the median and IQR.

#### Distribution of stillbirths by type

the number of stillbirths reported for each type divided by the total number of stillbirths per 100. This calculation was repeated for livebirths and neonatal deaths (death during the first 28 days of life following a livebirth) and the distributions compared.

#### Type specific Stillbirth Rate

Stillbirth rates for each type were calculated as **t**he number of stillbirths in the group divided by the total number of births in that group expressed as stillbirths per 1,000 total births (e.g., number of stillbirths between PT+SGA divided by number of total births between PT+SGA per 1000).

#### Stillbirth type specific Rate ratio

Rate ratios were calculated as the stillbirth rate in each type group, divided by the stillbirth rate in the reference group (T+AGA). These were calculated for each ‘newborn type’ and also for preterm types combined.

#### Population Attributable Fraction (PAF)

The prevalence of SGA type multiplied by the rate ratio in each type divided by the sum of the prevalence of SGA types multiplied by the rate ratio of all ‘newborn types’ in the population. We calculated PAF only for SGA types, as a proxy for fetal growth restriction, as fetal growth restriction is a potential pathway to stillbirth.

### Sensitivity Analysis

In view of the WHO recommendation for the use of late gestation stillbirth (≥28^+0^ weeks) for international comparisons and the potential large variations in ascertainment capture and reporting of early gestation stillbirth (22^+0^ – 27^+6^ weeks), we carried out a sensitivity analysis to explore if the distribution of stillbirth and stillbirth rate ratios differed if only late gestation stillbirths (28^+0^ to 44^+0^ weeks) were included.

## RESULTS

### Data quality assessment

Data were assessed from 14 national datasets collected between 2000 to 2020. We excluded country-years with >=20% missing birthweight or gestational age (Lebanon in: 2000-2019, Uruguay in: 2000-2019 and Malaysia in: 2010); missing information on livebirths (Argentina in 2019), and those which lay outside the study period (Iran in: 2021) (Figure 1a). Overall, 19.7.4% (29/147) and 9.5% (14/147) of country-years had >=20% missing birthweight data and missing gestational age, respectively and were excluded (Table S1b).

Data from 12 countries representing 119,644,788 total births (119,039,231 livebirths and 605,557 stillbirths) were included. Of the stillbirths, 218,351 were early gestation (22^+0^ - 27^+6^ weeks) and 387,206 late gestation (≥28^+0^ weeks). Data from a wide geographical range of high-income and middle-income countries were included (Figure 1b).

The overall stillbirth rate was 5.1 per 1,000 total births, with the highest rates in Iran, Mexico and Argentina (7.9, 7.8, and 7.4 per 1,000 total births respectively). The lowest stillbirth rate was observed in Sweden with 2.9 per 1,000 total births.

### Distribution of stillbirths by newborn type

The distribution of stillbirths according to the six ‘newborn types’ showed that most stillbirths (74.3%) were preterm types (PT+SGA (16.0%), PT+AGA (48.2%), PT+LGA (10.1%)) (Table 1a + Figure 2a). Under a fifth of stillbirths were T+AGA (16.7%), with around one in twenty T+SGA and T+LGA (5.0% and 4.2% respectively) (Table 1a).

**Table 1a.**
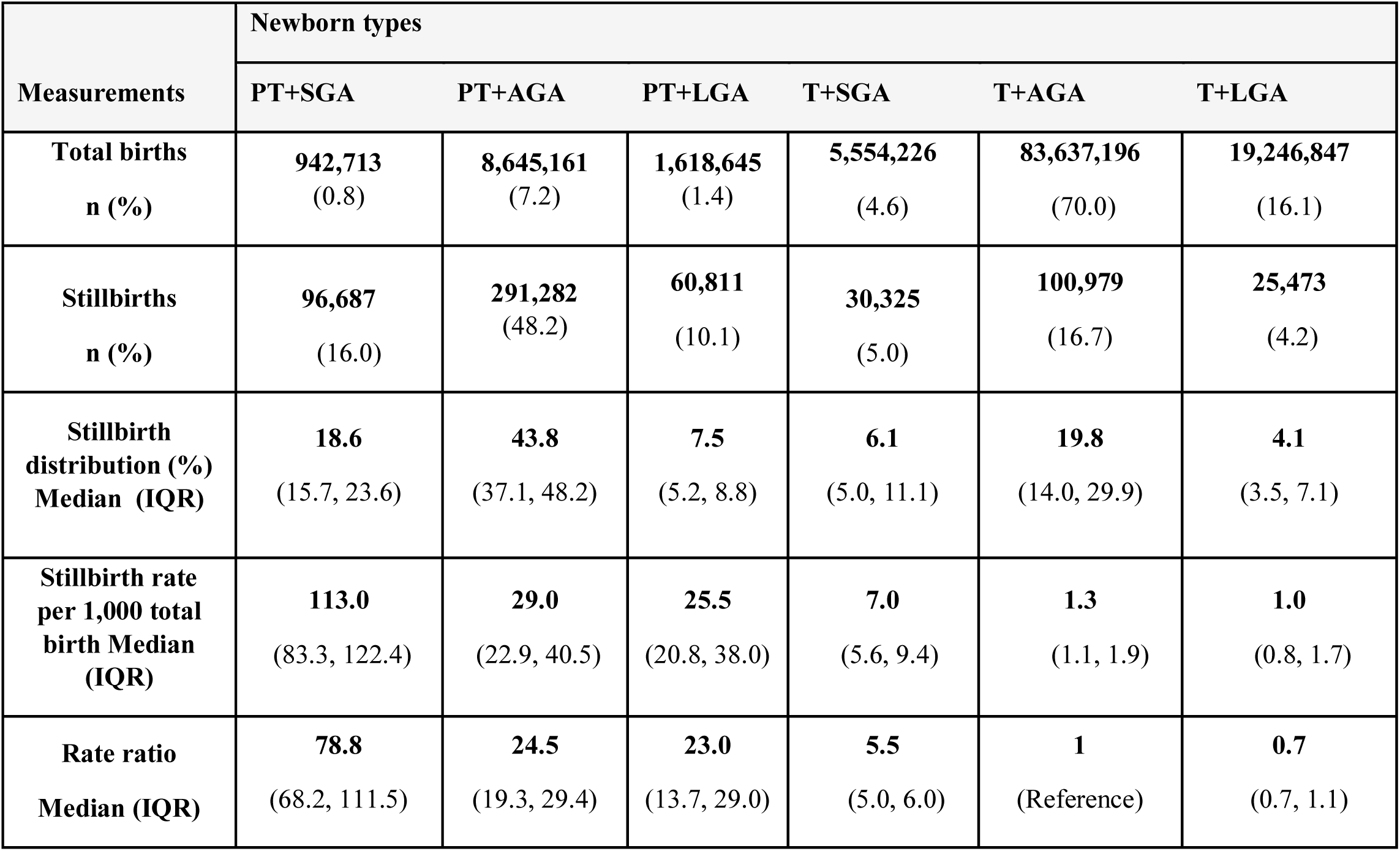
Stillbirth rate and rate ratio by newborn type for all stillbirths (≥22^+0^ weeks), 2000-2020.

| Measurements | Newborn types |  |  |  |  |  |
| --- | --- | --- | --- | --- | --- | --- |
|  | PT+SGA | PT+AGA | PT+LGA | T+SGA | T+AGA | T+LGA |
| **Total births**<br>n (%) | **942,713**<br>(0.8) | **8,645,161**<br>(7.2) | **1,618,645**<br>(1.4) | **5,554,226**<br>(4.6) | **83,637,196**<br>(70.0) | **19,246,847**<br>(16.1) |
| **Stillbirths**<br>n (%) | **96,687**<br>(16.0) | **291,282**<br>(48.2) | **60,811**<br>(10.1) | **30,325**<br>(5.0) | **100,979**<br>(16.7) | **25,473**<br>(4.2) |
| **Stillbirth distribution (%)**<br>**Median (IQR)** | **18.6**<br>(15.7, 23.6) | **43.8**<br>(37.1, 48.2) | **7.5**<br>(5.2, 8.8) | **6.1**<br>(5.0, 11.1) | **19.8**<br>(14.0, 29.9) | **4.1**<br>(3.5, 7.1) |
| **Stillbirth rate per 1,000 total birth Median (IQR)** | **113.0**<br>(83.3, 122.4) | **29.0**<br>(22.9, 40.5) | **25.5**<br>(20.8, 38.0) | **7.0**<br>(5.6, 9.4) | **1.3**<br>(1.1, 1.9) | **1.0**<br>(0.8, 1.7) |
| **Rate ratio**<br>**Median (IQR)** | **78.8**<br>(68.2, 111.5) | **24.5**<br>(19.3, 29.4) | **23.0**<br>(13.7, 29.0) | **5.5**<br>(5.0, 6.0) | **1**<br>(Reference) | **0.7**<br>(0.7, 1.1) |

**Table 1b.**
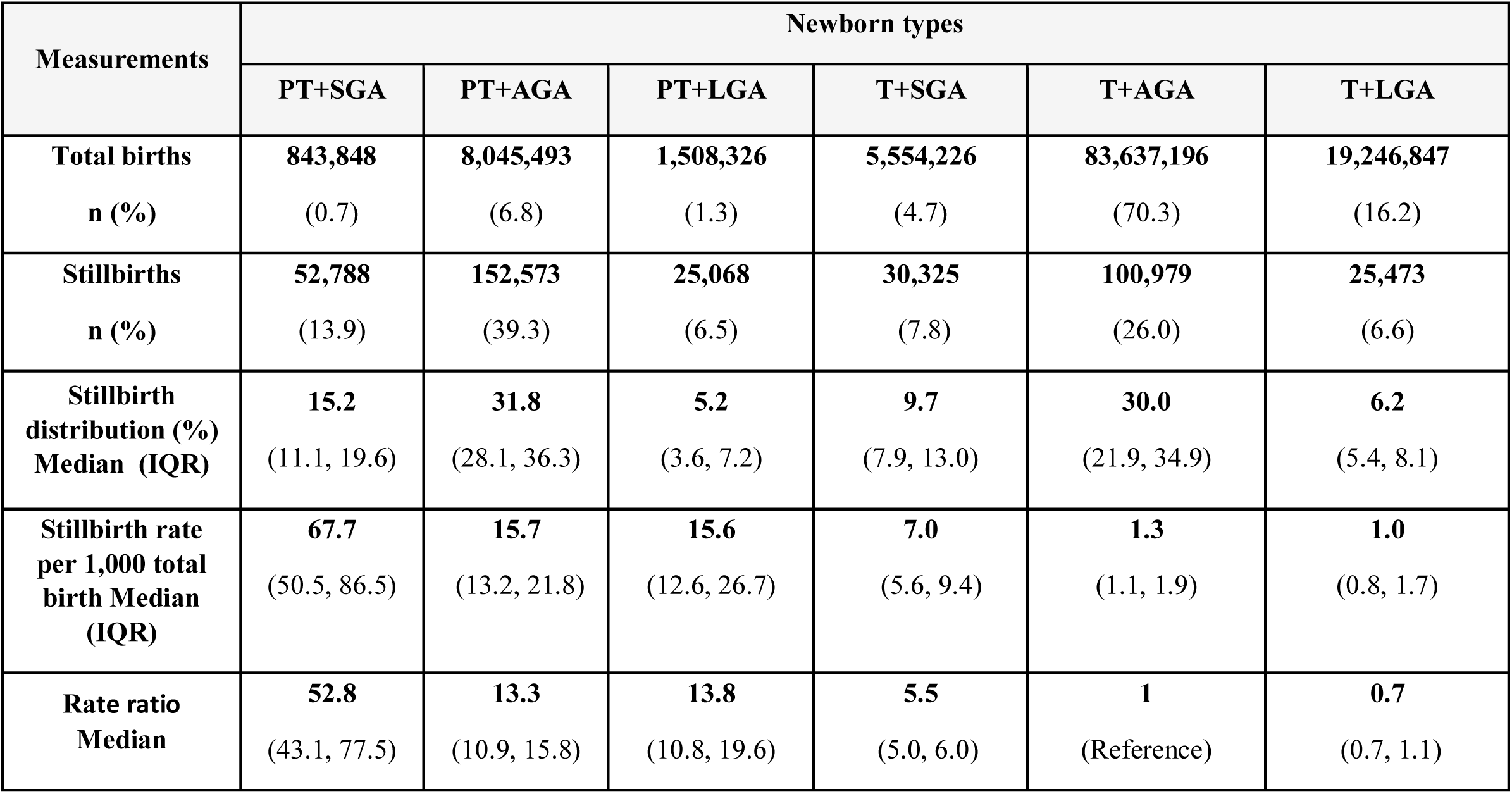
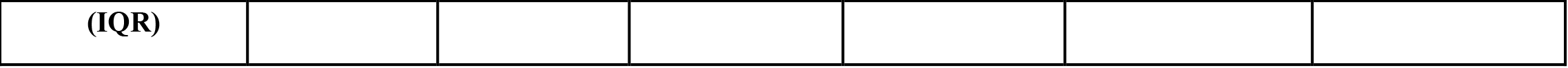
Stillbirth rate and rate ratio by newborn type for all stillbirths (≥28^+0^ weeks), 2000-2020.

**Figure 2.**
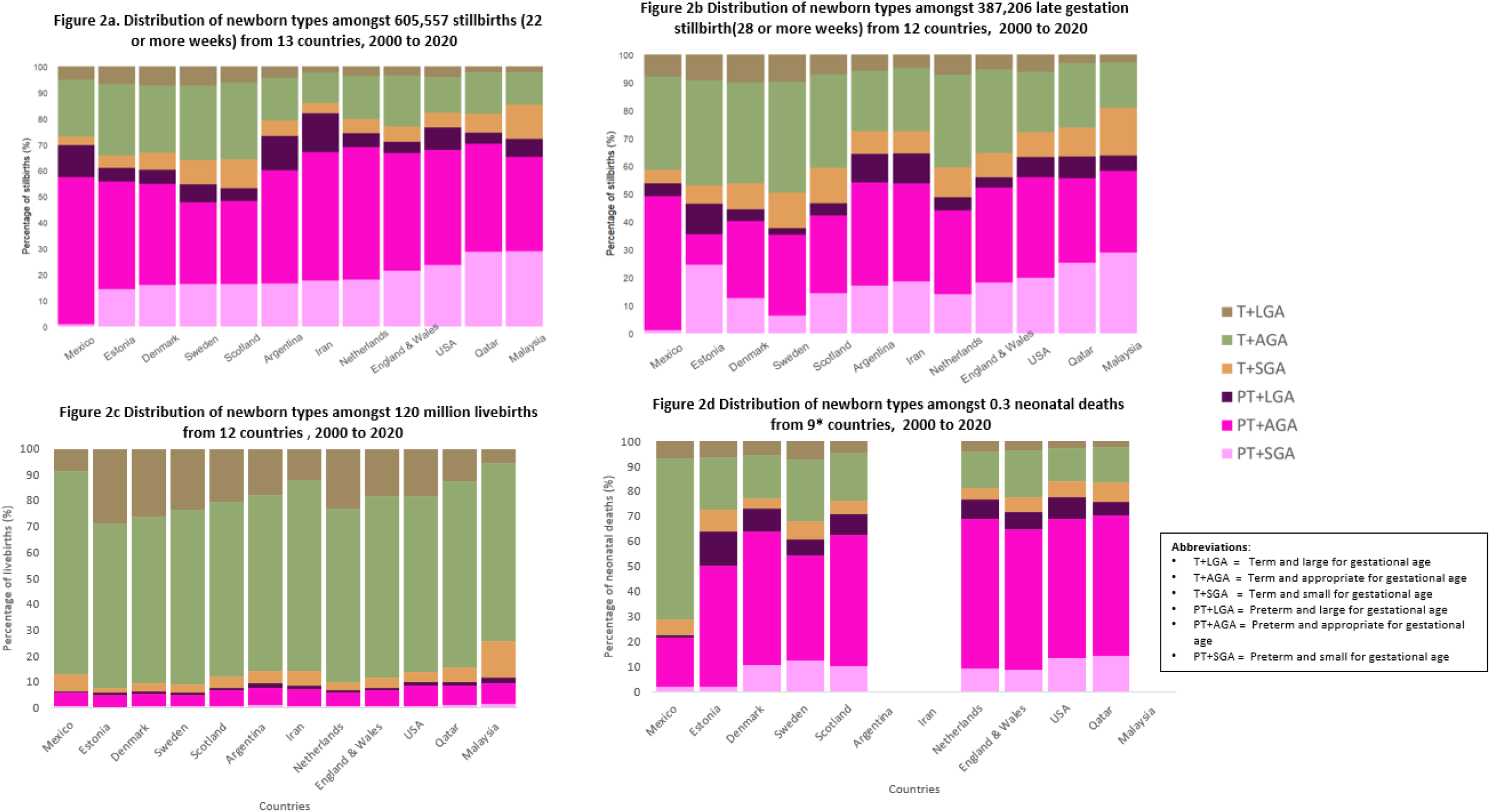
Distribution of ‘newborn types’ amongst a. all stillbirths^1^, b. late gestation stillbirths^2^, c. livebirths, d. neonatal deaths, 2000 to 2020. ^1^ See table S4a for further details. ^2^ See table S4b for further details.

There was substantial country level variation in the distribution of ‘newborn types’ among stillbirths. Overall, amongst all stillbirths ≥22^+0^ weeks, the median PT+SGA was 18.6% (IQR: 15.7 – 23.6) (ranging from 0.9% in Mexico to 28.8% in Malaysia); median PT+AGA 43.8% (IQR:37.1-48.2) (ranging from 31.1% in Sweden to 56.4% in Mexico); median PT+LGA 7.5% (IQR:5.2-8.8) (ranging from 4.4% in Qatar and England & Wales to 14.8% in Iran); median T+SGA 6.1% (IQR:5.0-11.1) (ranging from 3.2% in Mexico to 13.1% in Malaysia); median T+AGA 19.8% (IQR:14.0-29.9) (ranging from 11.6% in Iran to 92.5% in Scotland); median T+LGA 4.1% (IQR:3.5-7.1) (ranging from 2.0% in Qatar to 7.2% in Denmark) (Table 1a and Table S4a). Almost half of all stillbirths were preterm and AGA, with the highest percentages in Mexico 56.4% followed by the Netherlands (50.9), Iran (49.5%), England & Wales (45.3%) and USA (44.2%). Malaysia reported the highest prevalence of preterm and SGA stillbirth (28.9%), followed by Qatar (28.0%) and USA (23.7%). In contrast, Denmark, Sweden and Scotland reported relatively high percentages of term and LGA stillbirth, 7.2%, 7.1% and 5.9% respectively (Figure 2a, and Table S4a).

#### Comparison to distribution of ‘newborn types’ for livebirths and neonatal deaths

A similar pattern to the distribution of ‘newborn types’ for stillbirths was observed for neonatal deaths. Around 75% of neonatal deaths in all countries, apart from Mexico were preterm (Figure 2c). In contrast, most livebirths (91%) were born at term (T+AGA (70.2%), T+LGA (16.1%), T+SGA (4.6%)), with the remaining 10% preterm (PT+SGA (0.7%), PT+AGA (7.0%), PT+LGA (1.3%)) (Figure 2d).

### Rates of stillbirth by type

The overall stillbirth rate (including all stillbirths ≥22^+0^ weeks) for the study period was 5.1 per 1,000 total births. Stillbirth rates were highest for preterm ‘newborn types’: PT+SGA (median: 113.0 stillbirths per 1,000 total births, interquartile range IQR:83.3, 122.4), PT+AGA (median: 29.0, IQR:22.9, 40.5), and PT+LGA (median: 25.5, IQR:20.8, 38.0), followed by those T+SGA (median: 7.0 IQR:5.6, 9.4), T+AGA (median: 1.3 IQR:1.1, 1.9) and T+LGA (median: 1.0 IQR: 0.8, 1.7) (Table 1a). At country-level, the highest stillbirth rates among the PT+SGA types were observed in Iran (SBR: 149.2, 95%CI:149.0, 1149.4) and Qatar (SBR: 132.3, 95%CI: 131.4, 133.2) (Table S4a). Mexico, Iran, and Argentina had the highest three stillbirth rates in the PT+AGA types (SBR:77.3, 95%CI:77.2,77.4), (SBR: 57.2, 95%CI:57.0, 57.5), and (SBR: 44.9, 95%CI:44.9, 44.9) respectively) (Table S4a).

### Stillbirth rate ratios by ‘newborn type’

Compared with T+AGA, the median stillbirth rate ratio was around 80-fold (median RR: 78.8, IQR:68.2,111.5) for babies with the coexistence of preterm and SGA, over 20-fold for those PT+LGA (median RR: 25.5, IQR:20.8, 38.0) or PT+AGA (median RR: 29.0, IQR:22.9, 40.5), and 7-fold for babies T+SGA (median RR: 7.0, IQR:5.6, 9.4) (Table 1a, Figure 3a).

**Figure 3a.**
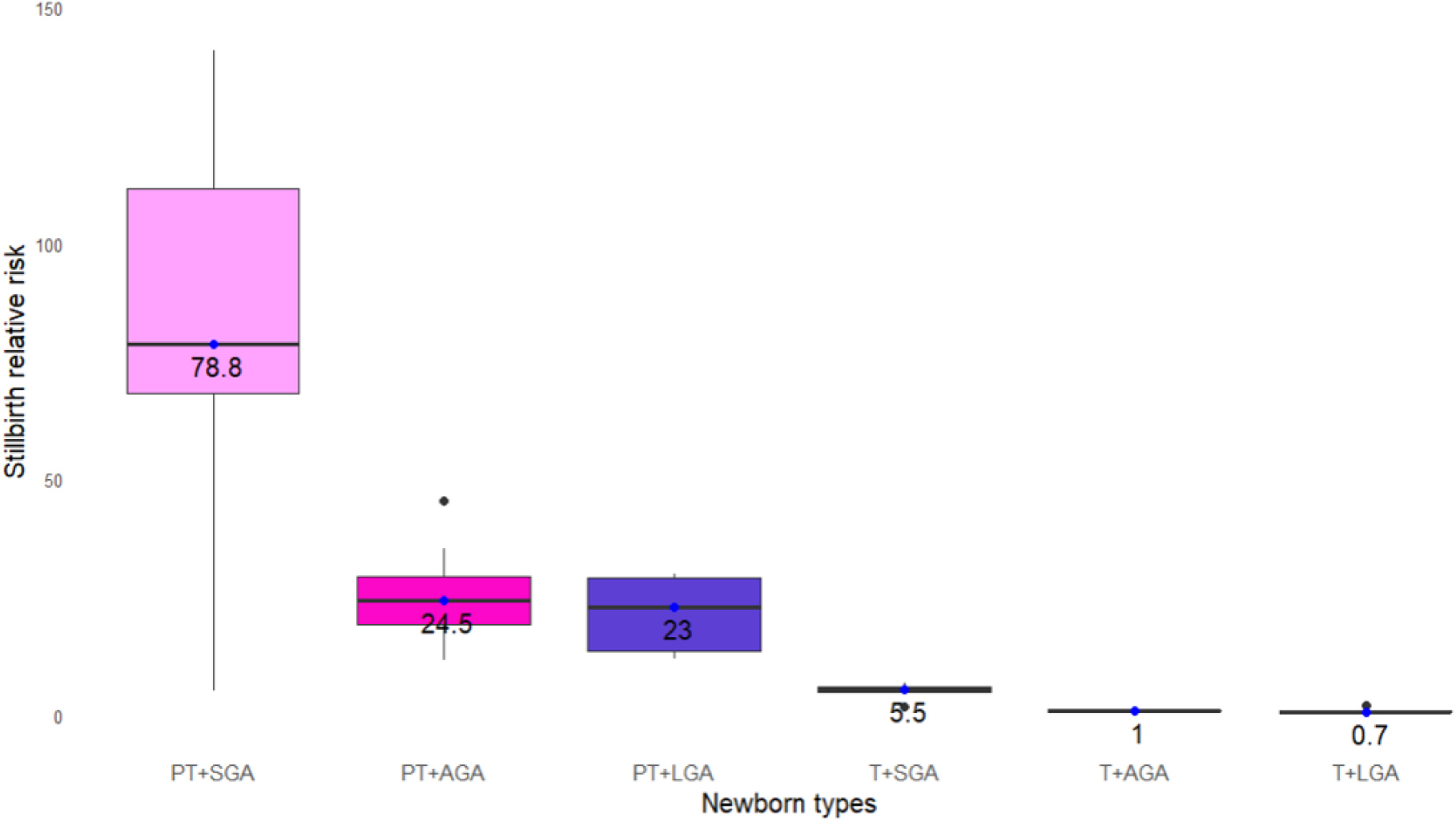
Stillbirth rate ratio by ‘newborn types’ amongst all stillbirths (≥22^+0^ weeks), 2000-2020. *Each point represents the stillbirth rate ratio, box plots summarise median values and IQR (25*^*th*^ *and 75*^*th*^ *percentiles). (countries=12, n=605,557) See Table S4a for further details)*.

**Figure 3b.**
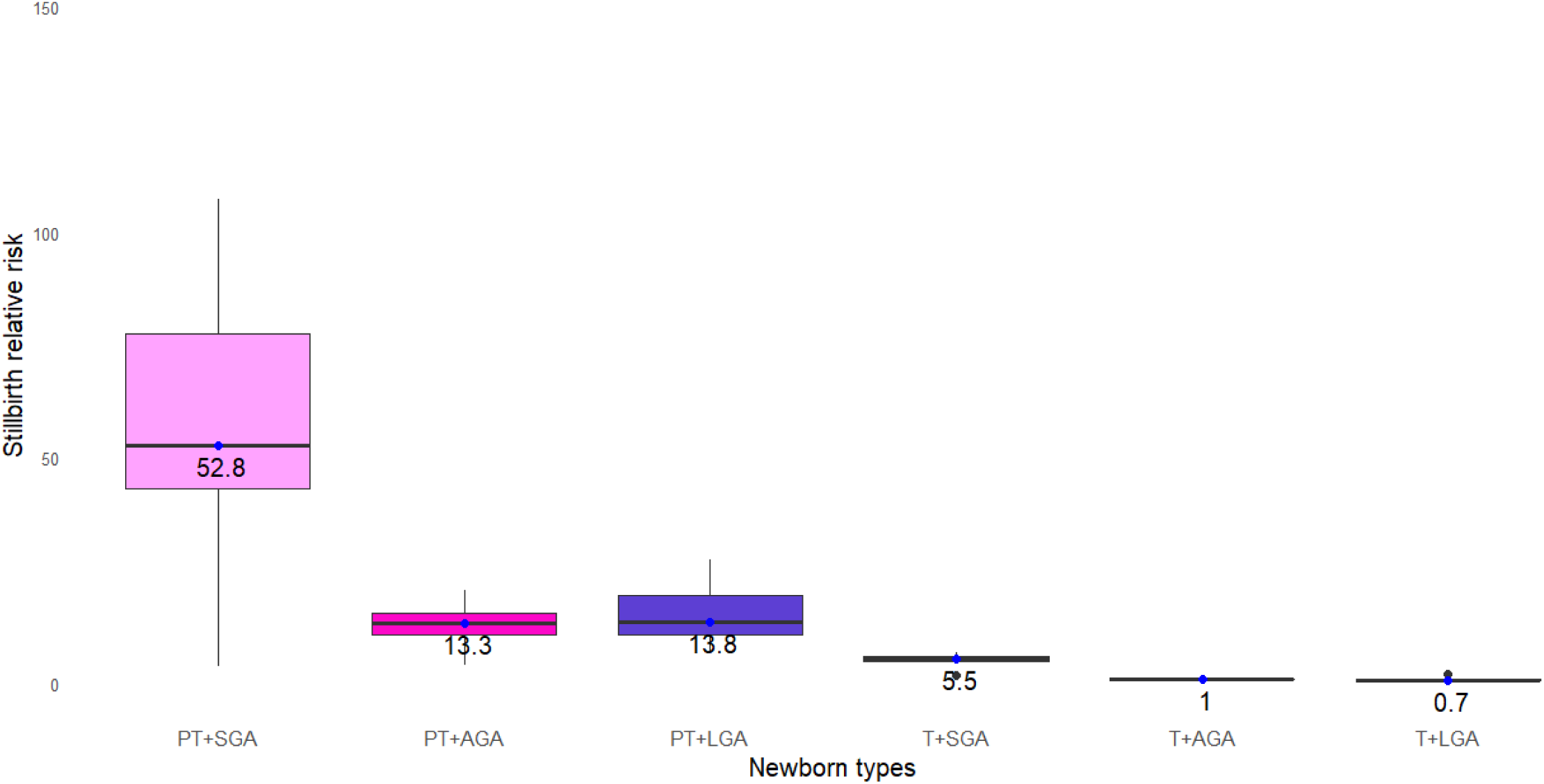
Stillbirth rate ratio by ‘newborn types’ amongst late gestation stillbirth (≥28^+0^ weeks), 2000-2020. *Each point represents the stillbirth rate ratio, box plots summarise median values and IQR (25*^*th*^ *and 75*^*th*^ *percentiles). (countries=12, n=387,206) See Table S4b for further details)*.

At country-level, the highest stillbirth rate ratio for PT+SGA was observed in USA (RR: 140.9, 95%CI [140.8, 141.0]), followed Qatar (RR: 117.4, 95%CI:116.6, 118.1) (Table S4a). Iran, Netherlands, and Mexico had the highest stillbirth rate ratios for PT+AGA types (RR: 45.4, 95%CI, 45.3, 45.5; RR: 35.6., 95%CI, 35.2, 36.1; RR:35.3, 95%CI (35.1, 35.5) respectively), Table S4a.

### Contribution of SGA to stillbirths (Population Attributable Fraction (PAF))

At the population level, over a fifth (21.5%) of stillbirths were attributable to being SGA (used as a proxy for growth-restriction) before term (PT+SGA median PAF: 19.0, IQR, 16.7, 26.0) with an additional 1.7% attributable to SGA at term (T+SGA median PAF:1.7, IQR, 1.2, 3.0).

### Sensitivity analyses

Preterm types remained the dominant type even when only late gestation stillbirths were included with around half of all stillbirths being preterm (Table 1b and Figure 2b).

A similar pattern in stillbirth rate and rate ratios were observed when only late gestation stillbirths were included, although the late gestation preterm ‘newborn type’ specific stillbirth rates were around two thirds of those for all births from 22^+0^ weeks and the stillbirth rate ratios for the PT+AGA and PT+LGA and half of those for all births from 22^+0^ weeks (Table 1b).

## DISCUSSION

### Main findings

This paper, including analyses of more than half a million babies from 12 countries stillborn between 2000 and 2020, has provided the first multi-country description of stillbirths using this novel classification by ‘newborn type’ combining attained size-for-gestational age and preterm or term. This classification goes beyond the traditional cut-offs and enables assessment of the contribution of preterm, SGA, and their combination. This has been shown to be useful for livebirths for identifying risk of neonatal death.^15,19^ Our results showed the overlap between preterm birth and stillbirth, with around three quarter of all stillbirths in these settings born preterm, compared with just 9.0% of livebirths. A fifth (21.0%) of stillbirths were SGA at birth, a substantially higher proportion than for livebirths (5.4%). The combination of preterm and SGA had the highest the stillbirth rate ratios compared with T+AGA. No additional stillbirth risk was found for term LGA babies compared with term AGA babies.

### Interpretation

Stillbirths are strongly associated with gestational ages <37^+0^ weeks.^20^ In this study, around 75% of stillbirths were preterm, slightly higher than that reported in a recent study in six low and middle -income countries, which reported 60% of stillbirths were preterm.^21^

We found the largest difference in stillbirth rates compared with T+AGA in all countries were for births that were both preterm and SGA (as a proxy of being growth restricted), followed by those PT+AGA, or PT+LGA. The increased risk for PT+LGA compared to appropriately grown term births is likely to be driven by low gestational age, rather than large size for gestational age. Overall stillbirth rates for preterm types were around 25 times higher than for term types. Consistent with previous research, this study found that those SGA at term were more likely to be stillborn than their appropriately grown peers.^22^

At population level, SGA (diagnosed at birth) contributed to around 21.5% of all stillbirths in these 12 countries with relatively high levels of pregnancy monitoring and interventional obstetrics, including provider-initiated delivery following in-utero diagnosis of severe fetal growth restriction. This is higher than the 11% population attributable risk reported in a previous study of eight high and middle-income countries.^23^ However, that study included only antepartum stillbirths from low-risk women and may not be generalisable to the whole population.

Understanding the population-level scale of the impact of fetal growth on stillbirth is crucial as stillbirths associated with fetal growth restriction are preventable with improved antenatal screening.^24^ However, there is a balance of risks between detecting fetal growth restriction and acting to prevent stillbirth, versus increasing preterm birth and associated complications.^25^ This balance of risks is even more pertinent in low-resource settings where full neonatal intensive care is less likely to be available. A recent multi-country study (Ghana, India, Kenya, Rwanda, and South Africa) found routine Doppler screening in a low-risk obstetric population an effective tool for reducing stillbirth rates.^26^ In France, antenatal detection of fetal growth restriction (FGR) was found to be protective against stillbirth, but despite detection of FGR, over 40% of stillbirths occurred among SGA babies.^27^

There is a major focus on small size at birth, however increasing evidence indicates that large for gestational age, which may be associated with the maternal metabolic environment, is also associated with an increased risk of stillbirth.^28,29^ In this study we found no increased risk of stillbirth in term babies who were LGA at birth compared to AGA, although this may be in part as the included populations had very low rates of post-term delivery where the risks associated with LGA may be greater. This finding differs from that of previous studies where the risk of stillbirth after 36^+0^ weeks gestation was higher for LGA compared to AGA pregnancies.^22,28,30^ However the use of INTERGROWTH-21^st^ newborn standard may have also accounted for these differences as it is known to left shift the centile distribution compared to national charts used in other studies.^31^

### Strength and limitations

A strength of our analyses is the large sample size combining data from across 12 countries with high data completeness and other measure of data quality. This has enabled exploration of associations with gestational age, by attained size for-gestational-age, and across time.

There are also limitations. Importantly, this study uses size-for-gestational age at birth as a proxy for fetal growth restriction (FGR). FGR is defined as the failure of the fetus to meet its growth potential due to a pathological factor, most commonly placental dysfunction.^24^ FGR is diagnosed by a drop of estimated fetal weight (EFW) centile on serial ultrasound measurement. In practice, this is not always available, and clinicians may rely on single ‘snapshot’ EFW assessment to define whether a baby is SGA in-utero – and are hence not able to differentiate whether an SGA baby is small due to pre-determined growth potential, or growth-faltering. In this study, the use of size-for-gestational age at birth rather than EFW in-utero may, in the rare cases where there is a prolonged period between fetal death and delivery, result in babies appropriately grown until the time of fetal death being classified at birth as SGA.

Secondly, to seek to provide comparability with live births, these ‘newborn types’ were based on the comparison to T+AGA. However, using a single dichotomous preterm versus term categorisation for stillbirths may not provide the level of granularity required, and importantly using such an approach it was not possible to estimate gestation-specific risk using a fetuses-at-risk approach.^32^

The comparability of results may be affected by the variation in gestational age assessment methods used (last menstrual period, different best obstetric estimates, ultrasound and the timing of ultrasound assessment) In addition our findings may also be affected by variations in stillbirth definition used by countries, and whether elective terminations of pregnancy are combined with stillbirths for reporting purposes (Table S5).^33^ It is well recognised that the reporting of births and misclassification between stillbirth and very early neonatal death is more common around the clinician’s perceptions of limits of viability and the thresholds of reporting in any given setting.^5,34,35^ Therefore shifting the threshold of reporting down to require reporting of all fetal deaths from 20^+0^ weeks will improve capture of all stillbirths from 22^+0^ weeks as defined by WHO.^1^ However, most countries only routinely recorded stillbirths from 22^+0^ weeks in their data system, with some only reporting from 24^+0^ weeks (Table S5). In the latter cases, whilst data were provided for this study on stillbirth at 22 or 23 weeks, there may be under capture as reporting of these deaths is not mandatory. Hence, we undertook a sensitivity analysis including only late gestation stillbirths at ≥28^+0^ weeks (63.4% of all stillbirths). This showed a similar pattern to the main analyses, with the highest rates and rate ratios for the preterm types, and as expected, the stillbirth rate and rate ratios by ‘newborn types’ were lower for all groups when considering only late gestation stillbirths compared to all stillbirths at ≥22^+0^ weeks.

Furthermore, despite around 98% of global stillbirths occurring in low-and middle-income countries, high-quality routine individual-level data on stillbirths from these countries are lacking and it was not possible to include these countries in this analysis.

Further research is required to assess the use of these ‘newborn types’ for stillbirths in higher burden contexts, especially those with high rates of SGA, notably South Asia.^14^ In addition, assessing risk by more detailed gestational age categories using a fetuses-at-risk approach, including data on labour-type (spontaneous vs provider-initiated), and combining these analyses with analyses of neonatal deaths could enable improved understanding of the epidemiology and provide data to target interventions, especially in settings with high levels of pregnancy monitoring and interventional obstetrics.^36^

## CONCLUSION

Our study provides the first multi-country analysis of ‘newborn types’ for stillbirths. Where individual level data are available categorisation of all births, including stillbirths, into these ‘newborn types’ was analytically possible using routinely collected data in these 12 upper-middle- or high-income contexts and led to programmatic relevant findings.

Preterm stillbirth accounted for more than three quarters of all stillbirths in these high-quality data from high/middle income countries. SGA is also associated with stillbirth, especially in combination with being preterm. More analyses of these ‘newborn types’ across a range of mortality contexts and extending gestation and size risk assessment using a fetuses-at-risk approach could provide more information on pathways to stillbirth and enable better targeting of interventions to underlying causes such as infections and obstetric complications.

## Data Availability

Data sharing and transfer agreements were jointly developed and signed by all collaborating partners. The pooled summary table data generated during the current study are deposited online with data access subject to approval at https://doi.org/10.17037/DATA.00003095 except for those from countries where data sharing is not permitted.

## Acknowledgements

Firstly, and most importantly, we thank all women and families included in national datasets, those who have led the data systems in these 12 countries, and all members of national teams. We thank all relevant national governments and other funders for their investments to enable the input data. We thank Claudia DaSilva and all the relevant administrative staff for their support.

## Authors’ contributions

The Vulnerable Newborn Measurement Collaboration was conceptualised by Joy Lawn and Bob Black. All collaborators contributed to the design of the study protocol. YO, HB, EOO, and JEL developed the detailed research questions and overall analysis plan for this paper. These were refined with inputs from the wider Vulnerable Newborn Measurement Collaboration. Analysis was undertaken by YO, LSI, HB and EOO provided statistical oversight. The manuscript was drafted by YO, LSI, and HB with EOO and JEL. All authors reviewed and agreed on the final version for publication.

## Ethics approval

The Vulnerable Newborn Measurement Collaboration was granted ethical approval from the Institutional Review Boards of the London School of Hygiene & Tropical Medicine (ref: 22858) and Johns Hopkins University. All the 12 country teams had ethical approval for use of data or exemptions based on current remit (Table S5).

## National Vulnerable Newborn Collaborative Group for Stillbirths

**Argentina** Veronica Pingray; Gabriela Gabriela; José Belizan; Luz Gibbons; Carlos Guevel

**Australia:** Vicki Flenady; Adrienne Gordon; Kara Warrilow; Harriet Lawford

**Denmark:** Erzsébet Horváth-Puhó, Henrik T Sørensen

**Estonia:** Luule Sakkeus; Liili Abuladze

**Lebanon:** Khalid A. Yunis; Ayah Al Bizri; Pascale Nakad

**Malaysia:** Shamala Karalasingam; J Ravichandran; R Jeganathan; Nurakman Binti Baharum

**Mexico:** Lorena Suárez-Idueta; Arturo Barranco Flores; Jesus Felipe Gonzalez Roldan; Sonia Lopez Alvarez

**the Netherlands:** Lisa Broeders; Aimée E. van Dijk

**Qatar:** Fawzia Alyafei; Mai AlQubaisi; Tawa O. Olukade; Hamdy A. Ali; Mohamad Rami Alturk

**Sweden:** Neda Razaz; Jonas Söderling

**UK_England and Wales:** Lucy K. Smith; Bradley N. Manktelow; Ruth J. Matthews; Elizabeth Draper; Alan Fenton; Jennifer J. Kurinczuk

**UK_Scotland:** Rachael Wood; Celina Davis; Kirsten Monteath; Samantha Clarke

**Uruguay:** Isabel Pereyra, Gabriella Pravia

**USA:** Sarka Lisonkova; Qi Wen

**LSHTM:** Joy E. Lawn; Hannah Blencowe; Eric Ohuma; Yemi Okwaraji; Judith Yargawa; Ellen Bradley

