## Supplementray figures and tables for "Stillbirths: contribution of preterm birth and size-for-gestational age for 119.6 million total births from nationwide records in 12 countries, 2000 to 2020"

Vulnerable Newborn multi-country analyses related to preterm births and small-for-gestational age.

### PAPER TITLE

### PAPER RUNNING TITLE

Stillbirth risk by newborn types in 12 countries.

### SUPPORTING INFORMATION

### S1: Definitions of stillbirth and input data

#### Table S1a. Definitions of stillbirths

**International Classification of Diseases (ICD-11)**

*Stillbirth:* A stillbirth is 'the complete expulsion or extraction from a woman of a fetus, following its death prior to the complete expulsion or extraction, at 22 or more completed weeks of gestation.

For international reporting it is recommended to report stillbirths of 28 or more completed weeks of gestation (late stillbirth). Countries with ability for reporting stillbirths of 22 or more completed weeks of gestation (early stillbirth) are recommended to do so.

When information on gestational age is unavailable for spontaneous abortion or stillbirth, use birthweight less than 500 grams as the criteria or use birthweight less than 1000 grams to report 28 or more completed weeks of gestation (late stillbirths) .

Sources:[1,2]

### Table S1b. Overview of input data to inform stillbirth analyses from13 countries, 2000-2020

| Country | Year | Live birth | Livebirth Missing values | | | | Livebirth Missing values | | | | Fetal deaths | Stillbirth Missing values | | | | Stillbirth Missing values | | | | Stillbirth |
|---|---|---|---|---|---|---|---|---|---|---|---|---|---|---|---|---|---|---|---|---|
| | | | BW | GA | BW & GA | Sex | <22 weeks | ≥ 45 weeks | > 6,500g | Implausible BW | | BW | GA | BW & GA | Sex | < 22 weeks | ≥45 weeks | > 6,500 g | Implausible BW | |
| Argentina | 2017 | 628,373 | 0.7 | 1.3 | 0.3 | 0.7 | 0.0 | 0.0 | 0.0 | 0.0 | 5,832 | 6.0 | 3.4 | 2.4 | 5.6 | 13.0 | 0.0 | 0.0 | 0.3 | 4,709 |
| Argentina | 2018 | 610,080 | 0.6 | 0.9 | 0.2 | 0.9 | 0.0 | 0.0 | 0.0 | 0.0 | 5,869 | 7.5 | 3.7 | 2.6 | 7.3 | 13.7 | 0.0 | 0.0 | 0.0 | 4,588 |
| Denmark | 2000-2013 | 867,290 | 1.9 | 1.7 | 1.5 | 0.0 | 0.0 | 0.0 | 0.0 | 0.0 | 3,723 | 16.6 | 2.6 | 2.4 | 13.7 | 0.6 | 0.0 | 0.0 | 0.0 | 2,991 |
| England & Wales | 2015 | 666,881 | 0.0 | 0.0 | 0.0 | 0.0 | 0.0 | 0.0 | 0.0 | 0.0 | 3,027 | 0.0 | 0.0 | 0.0 | 0.0 | 0.0 | 0.0 | 0.0 | 0.0 | 3,026 |
| England & Wales | 2016 | 666,539 | 0.0 | 0.0 | 0.0 | 0.0 | 0.0 | 0.0 | 0.0 | 0.0 | 3,009 | 0.0 | 0.0 | 0.0 | 0.0 | 0.0 | 0.0 | 0.0 | 0.0 | 3,009 |
| England & Wales | 2017 | 649,066 | 0.0 | 0.0 | 0.0 | 0.0 | 0.0 | 0.0 | 0.0 | 0.0 | 2,775 | 0.0 | 0.0 | 0.0 | 0.0 | 0.0 | 0.0 | 0.0 | 0.0 | 2,775 |
| England & Wales | 2018 | 621,468 | 0.0 | 0.0 | 0.0 | 0.0 | 0.0 | 0.0 | 0.0 | 0.0 | 2,580 | 0.0 | 0.0 | 0.0 | 0.0 | 0.0 | 0.0 | 0.0 | 0.0 | 2,580 |
| England & Wales | 2019 | 608,538 | 0.0 | 0.0 | 0.0 | 0.0 | 0.0 | 0.0 | 0.0 | 0.0 | 2,441 | 0.0 | 0.0 | 0.0 | 0.0 | 0.0 | 0.0 | 0.0 | 0.0 | 2,441 |
| Estonia | 2015 | 13,907 | 0.0 | 0.0 | 0.0 | 0.0 | 0.0 | 0.0 | 0.0 | 0.0 | 54 | 0.0 | 0.0 | 0.0 | 0.0 | 0.0 | 0.0 | 0.0 | 0.0 | 54 |
| Estonia | 2016 | 13,861 | 0.0 | 0.0 | 0.0 | 0.0 | 0.0 | 0.0 | 0.0 | 0.0 | 49 | 0.0 | 0.0 | 0.0 | 0.0 | 0.0 | 0.0 | 0.0 | 0.0 | 49 |
| Estonia | 2017 | 13,511 | 0.0 | 0.0 | 0.0 | 0.0 | 0.0 | 0.0 | 0.0 | 0.0 | 45 | 0.0 | 0.0 | 0.0 | 0.0 | 0.0 | 0.0 | 0.0 | 0.0 | 45 |
| Estonia | 2018 | 14,157 | 0.0 | 0.0 | 0.0 | 0.0 | 0.0 | 0.0 | 0.0 | 0.0 | 46 | 0.0 | 0.0 | 0.0 | 0.0 | 0.0 | 0.0 | 0.0 | 0.0 | 46 |
| Estonia | 2019 | 13,873 | 0.0 | 0.0 | 0.0 | 0.0 | 0.0 | 0.0 | 0.0 | 0.0 | 27 | 0.0 | 0.0 | 0.0 | 0.0 | 0.0 | 0.0 | 0.0 | 0.0 | 27 |
| Estonia | 2020 | 13,013 | 0.0 | 0.1 | 0.0 | 0.0 | 0.0 | 0.0 | 0.0 | 0.0 | 30 | 0.0 | 0.0 | 0.0 | 0.0 | 0.0 | 0.0 | 0.0 | 0.0 | 30 |
| Iran | 2017 | 1,170,242 | 0.0 | 0.0 | 0.0 | 0.0 | 0.0 | 0.0 | 0.0 | 0.1 | 8,882 | 0.0 | 0.0 | 0.0 | 0.0 | 0.0 | 0.0 | 0.1 | 0.5 | 8,734 |
| Iran | 2018 | 1,430,979 | 0.0 | 0.0 | 0.0 | 0.0 | 0.0 | 0.0 | 0.0 | 0.1 | 10,831 | 0.0 | 0.0 | 0.0 | 0.0 | 0.0 | 0.0 | 0.0 | 0.4 | 10,678 |
| Iran | 2019 | 1,274,706 | 0.0 | 0.0 | 0.0 | 0.0 | 0.0 | **0.0** | **0.0** | 0.1 | 9,604 | 0.2 | 0.1 | 0.1 | 0.1 | 0.0 | 0.0 | 0.0 | 0.4 | 9,451 |
| Iran | 2020 | 936,701 | 0.1 | 0.1 | 0.1 | 0.1 | 0.0 | 0.0 | 0.0 | 0.1 | 9,771 | 0.4 | 0.2 | 0.2 | 0.2 | 0.0 | 0.0 | 0.0 | 0.2 | 9,599 |
| Lebanon | 2003 | 6,787 | 1.4 | 1.8 | 0.6 | 1.8 | 0.0 | 0.0 | 0.0 | 0.0 | 23 | 56.5 | 8.7 | 4.3 | 8.7 | 21.7 | 0.0 | 0.0 | 0.0 | 8 |
| Lebanon | 2004 | 9,018 | 1.6 | 3.6 | 1.0 | 1.4 | 0.0 | 0.0 | 0.0 | 0.0 | 122 | 45.1 | 15.6 | 13.1 | 19.7 | 7.4 | 0.0 | 0.0 | 0.0 | 51 |
| Lebanon | 2005 | 12,606 | 1.5 | 2.4 | 0.6 | 3.5 | 0.0 | 0.0 | 0.0 | 0.0 | 125 | 46.4 | 7.2 | 4.8 | 10.4 | 11.2 | 0.0 | 0.0 | 0.0 | 54 |
| Lebanon | 2006 | 14,070 | 1.5 | 2.1 | 0.3 | 2.0 | 0.0 | 0.0 | 0.0 | 0.0 | 132 | 48.5 | 16.7 | 13.6 | 11.4 | 9.1 | 0.0 | 0.0 | 0.0 | 56 |
| Lebanon | 2007 | 14,872 | 2.4 | 2.1 | 0.6 | 3.8 | 0.0 | 0.0 | 0.0 | 0.0 | 109 | 62.4 | 6.4 | 5.5 | 11.0 | 4.6 | 0.0 | 0.0 | 0.0 | 37 |
| Lebanon | 2008 | 16,834 | 2.3 | 2.9 | 0.8 | 2.9 | 0.0 | 0.0 | 0.0 | 0.0 | 116 | 43.1 | 4.3 | 4.3 | 7.8 | 8.6 | 0.0 | 0.0 | 0.0 | 59 |
| Lebanon | 2009 | 18,600 | 3.5 | 7.0 | 2.7 | 3.8 | 0.0 | 0.0 | 0.0 | 0.0 | 171 | 36.8 | 11.7 | 4.7 | 13.5 | 11.7 | 0.0 | 0.0 | 0.0 | 81 |
| Lebanon | 2010 | 20,135 | 3.2 | 4.6 | 2.3 | 4.0 | 0.0 | 0.0 | 0.0 | 0.0 | 167 | 41.3 | 13.2 | 12.6 | 15.0 | 12.0 | 0.0 | 0.0 | 0.0 | 77 |
| Lebanon | 2011 | 20,770 | 4.5 | 5.9 | 3.2 | 4.7 | 0.0 | 0.0 | 0.0 | 0.0 | 131 | 26.0 | 6.1 | 5.3 | 6.9 | 14.5 | 0.0 | 0.0 | 0.0 | 75 |
| Lebanon | 2012 | 23,852 | 5.7 | 6.7 | 4.0 | 5.0 | 0.0 | 0.0 | 0.0 | 0.0 | 133 | 37.6 | 24.1 | 15.8 | 19.5 | 6.8 | 0.0 | 0.0 | 0.0 | 60 |
| Lebanon | 2013 | 25,022 | 6.1 | 7.0 | 4.7 | 5.6 | 0.0 | 0.0 | 0.0 | 0.0 | 176 | 20.5 | 8.0 | 5.7 | 10.8 | 18.2 | 0.0 | 0.0 | 0.6 | 102 |
| Lebanon | 2014 | 20,531 | 6.9 | 7.7 | 6.0 | 7.3 | 0.0 | 0.0 | 0.0 | 0.0 | 122 | 25.4 | 5.7 | 2.5 | 10.7 | 14.8 | 0.0 | 0.0 | 0.0 | 70 |

| Country | Year | Live birth | Livebirth Missing values | | | | Livebirth Missing values | | | | Fetal deaths | Stillbirth Missing values | | | | Stillbirth Missing values | | | | Stillbirth |
|---|---|---|---|---|---|---|---|---|---|---|---|---|---|---|---|---|---|---|---|---|
| | | | BW | GA | BW & GA | Sex | <22 weeks | ≥ 45 weeks | > 6,500g | Implausible BW | | BW | GA | BW & GA | Sex | < 22 weeks | ≥45 weeks | > 6,500 g | Implausible BW | |
| Lebanon | 2015 | 17,226 | 9.1 | 9.3 | 7.8 | 9.8 | 0.0 | 0.0 | 0.0 | 0.0 | 78 | 24.4 | 0.0 | 0.0 | 3.8 | 17.9 | 0.0 | 0.0 | 0.0 | 48 |
| Lebanon | 2016 | 16,916 | 6.5 | 6.7 | 5.3 | 7.6 | 0.0 | 0.0 | 0.0 | 0.0 | 66 | 22.7 | 1.5 | 0.0 | 7.6 | 24.2 | 0.0 | 0.0 | 0.0 | 39 |
| Lebanon | 2017 | 15,810 | 2.6 | 2.6 | 1.6 | 3.0 | 0.0 | 0.0 | 0.0 | 0.0 | 66 | 13.6 | 1.5 | 1.5 | 7.6 | 25.8 | 0.0 | 0.0 | 1.5 | 36 |
| Lebanon | 2018 | 16,167 | 2.7 | 2.2 | 1.6 | 2.8 | 0.0 | 0.0 | 0.0 | 0.0 | 77 | 26.0 | 5.2 | 5.2 | 9.1 | 15.6 | 0.0 | 0.0 | 1.3 | 44 |
| Lebanon | 2019 | 13,458 | 1.8 | 1.7 | 0.9 | 2.1 | 0.0 | 0.0 | 0.0 | 0.1 | 60 | 26.7 | 5.0 | 3.3 | 16.7 | 10.0 | 0.0 | 0.0 | 0.0 | 35 |
| Malaysia | 2011 | 94,788 | 1.0 | 36.4 | 0.5 | 1.4 | 0.0 | 0.0 | 0.0 | 0.2 | 974 | 2.1 | 17.2 | 0.5 | 2.0 | 1.0 | 0.1 | 0.3 | 0.2 | 731 |
| Malaysia | 2012 | 111,327 | 0.5 | 8.6 | 0.0 | 0.9 | 0.0 | 0.0 | 0.0 | 0.2 | 904 | 0.4 | 5.5 | 0.0 | 1.5 | 0.6 | 0.4 | 0.0 | 0.0 | 793 |
| Malaysia | 2013 | 111,444 | 0.2 | 4.6 | 0.0 | 0.0 | 0.0 | 0.0 | 0.0 | 0.3 | 886 | 0.5 | 7.9 | 0.0 | 1.0 | 0.8 | 0.1 | 0.2 | 0.2 | 752 |
| Malaysia | 2014 | 127,594 | 1.0 | 6.4 | 0.1 | 0.1 | 0.0 | 0.0 | 0.0 | 0.3 | 1,002 | 1.6 | 10.1 | 0.4 | 1.7 | 1.6 | 0.1 | 0.0 | 0.2 | 825 |
| Malaysia | 2015 | 132,064 | 1.8 | 8.8 | 0.1 | 0.1 | 0.0 | 0.0 | 0.0 | 0.3 | 1,080 | 2.9 | 9.4 | 0.3 | 1.6 | 1.9 | 0.0 | 0.4 | 0.5 | 875 |
| Malaysia | 2016 | 125,268 | 2.1 | 9.0 | 0.2 | 0.1 | 0.0 | 0.0 | 0.0 | 0.2 | 1,068 | 3.0 | 8.8 | 0.5 | 1.7 | 1.6 | 0.0 | 0.0 | 0.1 | 889 |
| Malaysia | 2017 | 112,193 | 0.4 | 32.7 | 0.1 | 0.0 | 0.0 | 0.0 | 0.0 | 0.2 | 1,209 | 1.4 | 0.9 | 0.0 | 1.2 | 1.3 | 0.0 | 0.0 | 0.1 | 1,100 |
| Mexico | 2008 | 1,835,062 | 6.7 | 1.1 | 0.1 | 0.2 | 0.0 | 0.0 | 0.0 | 0.0 | 17,397 | 0.0 | 0.0 | 0.0 | 1.4 | 0.0 | 0.0 | 0.0 | 0.0 | 17,155 |
| Mexico | 2009 | 1,913,409 | 6.7 | 0.9 | 0.1 | 0.1 | 0.0 | 0.0 | 0.0 | 0.0 | 16,992 | 0.0 | 0.0 | 0.0 | 1.5 | 0.0 | 0.0 | 0.0 | 0.0 | 16,742 |
| Mexico | 2010 | 1,931,096 | 6.7 | 0.6 | 0.1 | 0.1 | 0.0 | 0.0 | 0.0 | 0.0 | 16,732 | 0.0 | 0.0 | 0.0 | 1.4 | 0.0 | 0.0 | 0.0 | 0.0 | 16,492 |
| Mexico | 2011 | 2,024,743 | 6.5 | 0.5 | 0.1 | 0.1 | 0.0 | 0.0 | 0.0 | 0.0 | 16,575 | 0.0 | 0.0 | 0.0 | 1.4 | 0.0 | 0.0 | 0.0 | 0.0 | 16,337 |
| Mexico | 2012 | 2,067,890 | 6.3 | 0.4 | 0.1 | 0.1 | 0.0 | 0.0 | 0.0 | 0.0 | 15,922 | 0.0 | 0.0 | 0.0 | 1.3 | 0.0 | 0.0 | 0.0 | 0.0 | 15,710 |
| Mexico | 2013 | 2,056,273 | 6.4 | 0.3 | 0.0 | 0.1 | 0.0 | 0.0 | 0.0 | 0.0 | 15,449 | 0.0 | 0.0 | 0.0 | 1.4 | 0.0 | 0.0 | 0.0 | 0.0 | 15,234 |
| Mexico | 2014 | 2,043,653 | 6.3 | 0.2 | 0.0 | 0.1 | 0.0 | 0.0 | 0.0 | 0.0 | 15,100 | 0.0 | 0.0 | 0.0 | 1.6 | 0.0 | 0.0 | 0.0 | 0.0 | 14,860 |
| Mexico | 2015 | 2,019,716 | 6.0 | 0.2 | 0.0 | 0.1 | 0.0 | 0.0 | 0.0 | 0.0 | 15,198 | 0.8 | 0.0 | 0.0 | 2.0 | 0.0 | 0.0 | 0.0 | 0.0 | 14,783 |
| Mexico | 2016 | 1,962,380 | 5.8 | 0.1 | 0.0 | 0.1 | 0.0 | 0.0 | 0.0 | 0.0 | 14,439 | 0.8 | 0.0 | 0.0 | 1.7 | 0.0 | 0.0 | 0.0 | 0.0 | 14,078 |
| Mexico | 2017 | 1,947,994 | 5.8 | 0.1 | 0.0 | 0.1 | 0.0 | 0.0 | 0.0 | 0.0 | 14,575 | 1.0 | 0.0 | 0.0 | 1.8 | 0.0 | 0.0 | 0.0 | 0.0 | 14,182 |
| Mexico | 2018 | 1,848,682 | 4.7 | 0.2 | 0.0 | 0.1 | 0.0 | 0.0 | 0.0 | 0.0 | 15,412 | 1.1 | 0.0 | 0.0 | 1.8 | 0.0 | 0.0 | 0.0 | 0.0 | 14,965 |
| Mexico | 2019 | 1,763,891 | 5.7 | 0.2 | 0.0 | 0.1 | 0.0 | 0.0 | 0.0 | 0.0 | 15,151 | 0.2 | 0.0 | 0.0 | 1.9 | 0.0 | 0.0 | 0.0 | 0.0 | 14,842 |
| Netherlands | 2010 | 176,272 | 0.1 | 0.6 | 0.0 | 0.0 | 0.0 | 0.0 | 0.0 | 0.0 | 1,012 | 1.9 | 0.3 | 0.0 | 1.8 | 0.0 | 0.0 | 0.0 | 0.2 | 938 |
| Netherlands | 2011 | 174,276 | 0.1 | 0.6 | 0.0 | 0.0 | 0.0 | 0.0 | 0.0 | 0.0 | 971 | 2.3 | 0.5 | 0.0 | 1.6 | 0.0 | 0.0 | 0.0 | 0.2 | 896 |
| Netherlands | 2012 | 171,470 | 0.1 | 1.1 | 0.0 | 0.0 | 0.0 | 0.0 | 0.0 | 0.0 | 957 | 2.0 | 0.2 | 0.0 | 1.5 | 0.0 | 0.0 | 0.0 | 0.0 | 888 |
| Netherlands | 2013 | 166,799 | 0.1 | 0.7 | 0.0 | 0.0 | 0.0 | 0.0 | 0.0 | 0.0 | 863 | 3.9 | 0.7 | 0.0 | 2.2 | 0.0 | 0.0 | 0.0 | 0.0 | 759 |
| Netherlands | 2014 | 170,467 | 0.1 | 0.9 | 0.0 | 0.0 | 0.0 | 0.0 | 0.0 | 0.0 | 829 | 2.8 | 0.8 | 0.0 | 1.9 | 0.0 | 0.0 | 0.0 | 0.0 | 752 |
| Netherlands | 2015 | 165,454 | 0.1 | 0.7 | 0.0 | 0.0 | 0.0 | 0.0 | 0.0 | 0.0 | 862 | 3.6 | 0.9 | 0.0 | 2.1 | 0.0 | 0.0 | 0.0 | 0.1 | 774 |
| Netherlands | 2016 | 168,397 | 0.1 | 0.5 | 0.0 | 0.0 | 0.0 | 0.0 | 0.0 | 0.0 | 791 | 3.8 | 1.6 | 0.0 | 2.4 | 0.0 | 0.0 | 0.0 | 0.1 | 698 |
| Netherlands | 2017 | 164,952 | 0.2 | 0.7 | 0.0 | 0.0 | 0.0 | 0.0 | 0.0 | 0.0 | 819 | 3.2 | 0.5 | 0.0 | 3.1 | 0.0 | 0.0 | 0.0 | 0.0 | 744 |
| Netherlands | 2018 | 160,107 | 0.3 | 0.6 | 0.0 | 0.0 | 0.0 | 0.0 | 0.0 | 0.0 | 927 | 5.2 | 1.6 | 0.0 | 1.9 | 0.0 | 0.0 | 0.0 | 0.1 | 825 |

| Country | Year | Live birth | Livebirth Missing values | | | | Livebirth Missing values | | | | Fetal deaths | Stillbirth Missing values | | | | Stillbirth Missing values | | | | Stillbirth |
|---|---|---|---|---|---|---|---|---|---|---|---|---|---|---|---|---|---|---|---|---|
| | | | BW | GA | BW & GA | Sex | <22 weeks | ≥ 45 weeks | > 6,500g | Implausible BW | | BW | GA | BW & GA | Sex | < 22 weeks | ≥45 weeks | > 6,500 g | Implausible BW | |
| Netherlands | 2019 | 163,035 | 0.1 | 0.6 | 0.0 | 0.0 | 0.0 | 0.0 | 0.0 | 0.0 | 921 | 2.2 | 1.1 | 0.0 | 2.1 | 0.0 | 0.0 | 0.0 | 0.0 | 851 |
| Netherlands | 2020 | 163,536 | 0.1 | 0.4 | 0.0 | 0.0 | 0.0 | 0.0 | 0.0 | 0.0 | 942 | 2.8 | 0.2 | 0.0 | 1.6 | 0.0 | 0.0 | 0.0 | 0.2 | 872 |
| Qatar | 2016 | 22,035 | 0.3 | 1.9 | 0.0 | 0.0 | 0.0 | 0.0 | 0.0 | 0.0 | 63 | 3.2 | 0.0 | 0.0 | 4.8 | 4.8 | 0.0 | 0.0 | 0.0 | 55 |
| Qatar | 2017 | 23,936 | 0.4 | 1.2 | 0.0 | 0.0 | 0.0 | 0.0 | 0.0 | 0.0 | 202 | 3.0 | 0.0 | 0.0 | 2.5 | 6.9 | 0.0 | 0.0 | 0.0 | 171 |
| Qatar | 2018 | 23,549 | 0.3 | 1.0 | 0.0 | 0.0 | 0.0 | 0.0 | 0.0 | 0.0 | 170 | 4.1 | 0.6 | 0.0 | 4.1 | 3.5 | 0.0 | 0.0 | 1.8 | 149 |
| Qatar | 2019 | 24,817 | 0.4 | 0.7 | 0.0 | 0.0 | 0.0 | 0.0 | 0.0 | 0.0 | 152 | 5.3 | 2.0 | 0.0 | 5.3 | 5.3 | 0.0 | 0.0 | 0.0 | 140 |
| Scotland | 2000 | 52,462 | 0.0 | 0.0 | 0.0 | 0.0 | 0.0 | 0.0 | 0.0 | 0.0 | 321 | 13.7* | 0.3 | 0.0 | 15.3 | 0.0 | 0.0 | 0.0 | 0.0 | 270 |
| Scotland | 2001 | 51,247 | 0.0 | 0.0 | 0.0 | 0.0 | 0.0 | 0.0 | 0.0 | 0.0 | 338 | 12.7* | 0.0 | 0.0 | 13.9 | 0.0 | 0.0 | 0.0 | 0.0 | 284 |
| Scotland | 2002 | 50,399 | 0.0 | 0.0 | 0.0 | 0.0 | 0.0 | 0.0 | 0.0 | 0.0 | 308 | 14.0* | 0.0 | 0.0 | 14.6 | 0.0 | 0.0 | 0.0 | 0.0 | 257 |
| Scotland | 2003 | 51,515 | 0.0 | 0.0 | 0.0 | 0.0 | 0.0 | 0.0 | 0.0 | 0.0 | 330 | 16.1* | 0.0 | 0.0 | 16.7 | 0.0 | 0.0 | 0.0 | 0.0 | 267 |
| Scotland | 2004 | 53,044 | 0.0 | 0.0 | 0.0 | 0.0 | 0.0 | 0.0 | 0.0 | 0.0 | 350 | 10.9* | 0.0 | 0.0 | 12.6 | 0.0 | 0.0 | 0.0 | 0.0 | 300 |
| Scotland | 2005 | 52,756 | 0.1 | 0.0 | 0.0 | 0.0 | 0.0 | 0.0 | 0.0 | 0.0 | 310 | 11.9* | 0.0 | 0.0 | 15.2 | 0.0 | 0.0 | 0.0 | 0.0 | 257 |
| Scotland | 2006 | 53,417 | 0.1 | 0.0 | 0.0 | 0.0 | 0.0 | 0.0 | 0.0 | 0.0 | 314 | 10.2* | 0.0 | 0.0 | 11.1 | 0.0 | 0.0 | 0.0 | 0.6 | 266 |
| Scotland | 2007 | 56,395 | 0.1 | 0.1 | 0.0 | 0.0 | 0.0 | 0.0 | 0.0 | 0.0 | 363 | 10.7* | 0.0 | 0.0 | 13.8 | 0.0 | 0.0 | 0.0 | 0.0 | 308 |
| Scotland | 2008 | 58,543 | 0.1 | 0.1 | 0.0 | 0.0 | 0.0 | 0.0 | 0.0 | 0.0 | 344 | 9.6* | 0.3 | 0.3 | 12.5 | 0.0 | 0.0 | 0.0 | 0.0 | 298 |
| Scotland | 2009 | 57,739 | 0.1 | 0.1 | 0.0 | 0.0 | 0.0 | 0.0 | 0.0 | 0.0 | 361 | 14.1* | 0.0 | 0.0 | 14.1 | 0.0 | 0.0 | 0.0 | 0.6 | 300 |
| Scotland | 2010 | 57,795 | 0.0 | 0.0 | 0.0 | 0.0 | 0.0 | 0.0 | 0.0 | 0.0 | 310 | 13.5* | 0.0 | 0.0 | 14.2 | 0.0 | 0.0 | 0.0 | 0.0 | 262 |
| Scotland | 2011 | 57,447 | 0.0 | 0.1 | 0.0 | 0.0 | 0.0 | 0.0 | 0.0 | 0.0 | 324 | 11.7* | 0.0 | 0.0 | 12.0 | 0.0 | 0.0 | 0.0 | 0.0 | 280 |
| Scotland | 2012 | 56,766 | 0.1 | 0.1 | 0.0 | 0.0 | 0.0 | 0.0 | 0.0 | 0.0 | 297 | 15.5* | 0.0 | 0.0 | 15.2 | 0.0 | 0.0 | 0.0 | 0.0 | 248 |
| Scotland | 2013 | 54,870 | 0.2 | 0.1 | 0.0 | 0.0 | 0.0 | 0.0 | 0.0 | 0.0 | 259 | 15.4* | 0.0 | 0.0 | 14.3 | 0.0 | 0.0 | 0.4 | 0.4 | 216 |
| Scotland | 2014 | 55,562 | 0.1 | 0.4 | 0.0 | 0.0 | 0.0 | 0.0 | 0.0 | 0.0 | 251 | 11.6* | 0.4 | 0.4 | 12.4 | 0.0 | 0.0 | 0.0 | 0.0 | 218 |
| Scotland | 2015 | 54,095 | 0.1 | 0.4 | 0.0 | 0.0 | 0.0 | 0.0 | 0.0 | 0.0 | 225 | 12.0* | 0.0 | 0.0 | 12.4 | 0.0 | 0.0 | 0.0 | 0.0 | 193 |
| Scotland | 2016 | 53,242 | 0.2 | 0.8 | 0.0 | 0.0 | 0.0 | 0.0 | 0.0 | 0.0 | 249 | 13.7* | 0.0 | 0.0 | 13.3 | 0.0 | 0.0 | 0.0 | 0.0 | 212 |
| Scotland | 2017 | 51,643 | 0.5 | 0.5 | 0.0 | 0.0 | 0.0 | 0.0 | 0.0 | 0.0 | 229 | 10.9* | 0.0 | 0.0 | 10.5 | 0.0 | 0.0 | 0.0 | 0.0 | 202 |
| Scotland | 2018 | 50,317 | 0.4 | 0.0 | 0.0 | 0.1 | 0.0 | 0.0 | 0.0 | 0.0 | 204 | 17.2* | 0.0 | 0.0 | 16.7 | 0.0 | 0.0 | 0.0 | 0.0 | 167 |
| Scotland | 2019 | 48,609 | 0.1 | 0.0 | 0.0 | 0.0 | 0.0 | 0.0 | 0.0 | 0.0 | 184 | 12.5* | 0.0 | 0.0 | 13.6 | 0.0 | 0.0 | 0.0 | 0.0 | 154 |
| Scotland | 2020 | 46,641 | 0.1 | 0.0 | 0.0 | 0.0 | 0.0 | 0.0 | 0.0 | 0.0 | 190 | 5.3* | 0.0 | 0.0 | 5.3 | 0.0 | 0.0 | 0.0 | 0.0 | 168 |
| Sweden | 2008 | 105,724 | 3.8 | 0.0 | 0.9 | 0.1 | 0.0 | 7.0 | 0.0 | 5.7 | 382 | 2.9 | 0.0 | 0.0 | 0.0 | 0.0 | 2.6 | 0.0 | 10.2 | 315 |
| Sweden | 2009 | 107,171 | 0.0 | 0.0 | 0.0 | 0.0 | 0.0 | 0.0 | 0.0 | 0.0 | 438 | 2.7 | 0.0 | 0.0 | 0.0 | 0.0 | 2.1 | 0.0 | 11.2 | 335 |
| Sweden | 2010 | 112,571 | 0.0 | 0.0 | 0.0 | 0.0 | 0.0 | 0.0 | 0.0 | 0.0 | 420 | 4.0 | 0.0 | 0.0 | 0.0 | 0.0 | 0.0 | 0.0 | 9.0 | 330 |
| Sweden | 2011 | 109,002 | 0.0 | 0.0 | 0.0 | 0.0 | 0.0 | 0.0 | 0.0 | 0.0 | 432 | 3.7 | 0.0 | 0.0 | 0.0 | 0.0 | 0.0 | 0.0 | 10.4 | 337 |
| Sweden | 2012 | 109,908 | 0.0 | 0.0 | 0.0 | 0.0 | 0.0 | 0.0 | 0.0 | 0.0 | 428 | 3.0 | 0.0 | 0.0 | 0.0 | 0.0 | 1.9 | 0.0 | 8.9 | 344 |
| Sweden | 2013 | 110,383 | 0.0 | 0.0 | 0.0 | 0.0 | 0.0 | 0.0 | 0.0 | 0.0 | 416 | 3.8 | 0.0 | 0.0 | 0.0 | 0.0 | 1.9 | 0.0 | 10.8 | 328 |
| Sweden | 2014 | 112,856 | 0.0 | 0.0 | 0.0 | 0.0 | 0.0 | 0.0 | 0.0 | 0.0 | 450 | 2.7 | 0.0 | 0.0 | 0.0 | 0.0 | 1.6 | 0.0 | 9.1 | 360 |
| Sweden | 2015 | 113,267 | 0.0 | 0.0 | 0.0 | 0.0 | 0.0 | 0.0 | 0.0 | 0.0 | 418 | 5.7 | 0.0 | 0.0 | 0.0 | 0.0 | 3.1 | 0.0 | 13.9 | 317 |

| Country | Year | Live birth | Livebirth Missing values | | | | Livebirth Missing values | | | | Fetal deaths | Stillbirth Missing values | | | | Stillbirth Missing values | | | | Stillbirth |
|---|---|---|---|---|---|---|---|---|---|---|---|---|---|---|---|---|---|---|---|---|
| | | | BW | GA | BW & GA | Sex | <22 weeks | ≥ 45 weeks | > 6,500g | Implausible BW | | BW | GA | BW & GA | Sex | < 22 weeks | ≥45 weeks | > 6,500 g | Implausible BW | |
| Sweden | 2016 | 115,944 | 0.0 | 0.0 | 0.0 | 0.0 | 0.0 | 0.0 | 0.0 | 0.0 | 417 | 2.9 | 0.0 | 0.0 | 1.9 | 0.0 | 1.9 | 0.0 | 11.3 | 326 |
| Sweden | 2017 | 113,963 | 0.0 | 0.0 | 0.0 | 0.0 | 0.0 | 0.0 | 0.0 | 0.0 | 414 | 1.7 | 0.0 | 0.0 | 1.2 | 0.0 | 3.1 | 0.0 | 10.6 | 330 |
| Sweden | 2018 | 114,630 | 0.0 | 0.0 | 0.0 | 0.0 | 0.0 | 0.0 | 0.0 | 0.0 | 435 | 3.2 | 0.0 | 0.0 | 1.4 | 0.0 | 2.5 | 0.0 | 10.6 | 340 |
| Sweden | 2019 | 113,343 | 0.0 | 0.0 | 0.0 | 0.0 | 0.0 | 0.0 | 0.0 | 0.0 | 363 | 4.1 | 0.0 | 0.0 | 0.0 | 0.0 | 0.0 | 0.0 | 11.3 | 297 |
| Uruguay | 2009 | 39,938 | 0.1 | 2.4 | 0.1 | 0.0 | 0.2 | 0.0 | 0.0 | 0.1 | 1,242 | 56.6 | 66.9 | 53.1 | 63.0 | 4.8 | 0.0 | 0.0 | 0.0 | 336 |
| Uruguay | 2010 | 33,145 | 0.0 | 1.5 | 0.0 | 0.0 | 0.1 | 0.0 | 0.0 | 0.0 | 1,056 | 62.7 | 71.3 | 59.5 | 67.0 | 4.3 | 0.0 | 0.0 | 0.0 | 252 |
| Uruguay | 2011 | 41,174 | 0.1 | 1.4 | 0.1 | 0.0 | 0.1 | 0.0 | 0.0 | 0.1 | 640 | 40.9 | 52.7 | 39.4 | 43.4 | 3.3 | 0.0 | 0.0 | 0.0 | 273 |
| Uruguay | 2012 | 44,727 | 0.1 | 1.1 | 0.1 | 0.0 | 0.0 | 0.0 | 0.0 | 0.1 | 425 | 9.9 | 29.9 | 9.2 | 10.8 | 3.1 | 0.0 | 0.0 | 0.0 | 280 |
| Uruguay | 2013 | 45,900 | 0.2 | 0.9 | 0.2 | 0.0 | 0.0 | 0.0 | 0.0 | 0.1 | 708 | 49.7 | 64.7 | 49.4 | 50.4 | 0.4 | 0.0 | 0.0 | 0.0 | 243 |
| Uruguay | 2014 | 46,438 | 0.0 | 0.7 | 0.0 | 0.0 | 0.0 | 0.0 | 0.0 | 0.0 | 636 | 40.6 | 54.1 | 39.9 | 42.0 | 0.8 | 0.0 | 0.0 | 0.2 | 276 |
| Uruguay | 2015 | 46,475 | 0.2 | 0.8 | 0.1 | 0.0 | 0.0 | 0.0 | 0.0 | 0.0 | 490 | 43.5 | 58.6 | 42.4 | 45.9 | 1.4 | 0.0 | 0.0 | 0.0 | 191 |
| Uruguay | 2016 | 45,076 | 0.1 | 0.8 | 0.1 | 0.0 | 0.0 | 0.0 | 0.0 | 0.1 | 570 | 36.0 | 48.6 | 35.1 | 38.6 | 1.8 | 0.0 | 0.0 | 0.0 | 272 |
| Uruguay | 2017 | 41,609 | 0.1 | 0.6 | 0.1 | 0.0 | 0.0 | 0.0 | 0.0 | 0.0 | 448 | 37.7 | 48.9 | 36.6 | 39.7 | 1.1 | 0.0 | 0.0 | 0.0 | 215 |
| Uruguay | 2018 | 38,459 | 0.1 | 0.8 | 0.1 | 0.0 | 0.0 | 0.0 | 0.0 | 0.0 | 372 | 38.4 | 50.5 | 37.4 | 39.2 | 2.4 | 0.0 | 0.0 | 0.0 | 171 |
| Uruguay | 2019 | 35,341 | 0.1 | 0.9 | 0.1 | 0.0 | 0.0 | 0.0 | 0.0 | 0.0 | 324 | 27.8 | 39.2 | 27.2 | 29.0 | 1.5 | 0.0 | 0.0 | 0.0 | 187 |
| Uruguay | 2020 | 34,543 | 0.1 | 1.0 | 0.1 | 0.0 | 0.0 | 0.0 | 0.0 | 0.0 | 292 | 20.2 | 30.5 | 19.9 | 26.7 | 6.2 | 0.0 | 0.0 | 0.3 | 180 |
| USA | 2000 | 4,007,957 | 0.1 | 1.1 | 0.1 | 0.0 | 0.1 | 0.1 | 0.0 | 0.0 | 27,046 | 11.1 | 3.4 | 1.2 | 4.6 | 22.0 | 0.1 | 0.0 | 0.4 | 17,665 |
| USA | 2001 | 3,979,543 | 0.1 | 1.0 | 0.0 | 0.0 | 0.1 | 0.1 | 0.0 | 0.0 | 26,408 | 10.5 | 3.3 | 1.3 | 4.2 | 21.6 | 0.1 | 0.0 | 0.3 | 17,477 |
| USA | 2002 | 3,974,227 | 0.1 | 1.0 | 0.0 | 0.0 | 0.1 | 0.1 | 0.0 | 0.0 | 25,980 | 10.5 | 3.0 | 1.1 | 4.2 | 22.0 | 0.1 | 0.0 | 0.3 | 17,072 |
| USA | 2003 | 4,039,360 | 0.1 | 1.1 | 0.0 | 0.0 | 0.1 | 0.1 | 0.0 | 0.0 | 25,683 | 9.7 | 1.4 | 0.0 | 4.0 | 22.8 | 0.1 | 0.0 | 0.3 | 17,018 |
| USA | 2004 | 4,062,714 | 0.1 | 1.1 | 0.0 | 0.0 | 0.1 | 0.1 | 0.0 | 0.0 | 25,706 | 10.2 | 1.3 | 0.0 | 4.0 | 22.5 | 0.1 | 0.0 | 0.3 | 17,158 |
| USA | 2005 | 4,102,847 | 0.1 | 0.7 | 0.0 | 0.0 | 0.1 | 0.1 | 0.0 | 0.0 | 25,931 | 11.9 | 2.5 | 1.0 | 4.2 | 22.8 | 0.1 | 0.0 | 0.2 | 16,817 |
| USA | 2006 | 4,234,499 | 0.1 | 0.6 | 0.0 | 0.0 | 0.1 | 0.1 | 0.0 | 0.0 | 26,002 | 12.0 | 2.0 | 0.5 | 4.6 | 22.2 | 0.1 | 0.0 | 0.3 | 17,268 |
| USA | 2007 | 4,307,092 | 0.1 | 0.2 | 0.0 | 0.0 | 0.1 | 0.0 | 0.0 | 0.0 | 26,632 | 12.4 | 1.0 | 0.4 | 4.8 | 23.8 | 0.0 | 0.0 | 0.3 | 17,533 |
| USA | 2008 | 4,240,158 | 0.1 | 0.1 | 0.0 | 0.0 | 0.1 | 0.0 | 0.0 | 0.0 | 26,367 | 13.0 | 0.7 | 0.3 | 4.6 | 23.3 | 0.0 | 0.0 | 0.3 | 17,600 |
| USA | 2009 | 4,124,198 | 0.1 | 0.1 | 0.0 | 0.0 | 0.1 | 0.0 | 0.0 | 0.0 | 24,902 | 9.4 | 0.6 | 0.2 | 4.7 | 23.4 | 0.0 | 0.0 | 0.3 | 16,588 |
| USA | 2010 | 3,993,492 | 0.1 | 0.1 | 0.0 | 0.0 | 0.1 | 0.0 | 0.0 | 0.0 | 24,276 | 9.4 | 0.7 | 0.2 | 4.7 | 23.3 | 0.0 | 0.0 | 0.3 | 16,151 |
| USA | 2011 | 3,947,847 | 0.1 | 0.1 | 0.0 | 0.0 | 0.1 | 0.0 | 0.0 | 0.0 | 24,319 | 9.5 | 0.6 | 0.2 | 4.9 | 23.8 | 0.0 | 0.0 | 0.3 | 16,193 |
| USA | 2012 | 3,947,771 | 0.1 | 0.1 | 0.0 | 0.0 | 0.1 | 0.0 | 0.0 | 0.0 | 24,108 | 9.3 | 0.7 | 0.3 | 4.9 | 23.4 | 0.0 | 0.0 | 0.2 | 15,924 |
| USA | 2013 | 3,927,847 | 0.1 | 0.1 | 0.0 | 0.0 | 0.1 | 0.0 | 0.0 | 0.0 | 23,621 | 9.1 | 0.5 | 0.2 | 4.7 | 23.0 | 0.0 | 0.0 | 0.2 | 15,854 |
| USA | 2014 | 3,987,017 | 0.1 | 0.1 | 0.0 | 0.0 | 0.1 | 0.0 | 0.0 | 0.0 | 24,032 | 7.7 | 0.6 | 0.2 | 3.4 | 21.8 | 0.0 | 0.0 | 0.2 | 16,246 |
| USA | 2015 | 3,977,767 | 0.1 | 0.1 | 0.0 | 0.0 | 0.1 | 0.0 | 0.0 | 0.0 | 23,811 | 7.3 | 0.5 | 0.1 | 3.2 | 21.9 | 0.0 | 0.0 | 0.2 | 16,256 |
| USA | 2016 | 3,944,206 | 0.1 | 0.1 | 0.0 | 0.0 | 0.1 | 0.0 | 0.0 | 0.0 | 23,921 | 6.8 | 0.5 | 0.1 | 3.1 | 22.4 | 0.0 | 0.0 | 0.2 | 16,263 |
| USA | 2017 | 3,855,137 | 0.1 | 0.1 | 0.0 | 0.0 | 0.1 | 0.0 | 0.0 | 0.0 | 22,860 | 7.2 | 0.6 | 0.1 | 3.4 | 21.8 | 0.0 | 0.0 | 0.1 | 15,637 |
| USA | 2018 | 3,792,323 | 0.1 | 0.1 | 0.0 | 0.0 | 0.1 | 0.0 | 0.0 | 0.0 | 22,540 | 7.1 | 0.5 | 0.2 | 3.2 | 22.3 | 0.0 | 0.0 | 0.2 | 15,338 |

| Country | Year | Live birth | Livebirth Missing values | | | | Livebirth Missing values | | | | Fetal deaths | Stillbirth Missing values | | | | Stillbirth Missing values | | | | Stillbirth |
|---|---|---|---|---|---|---|---|---|---|---|---|---|---|---|---|---|---|---|---|---|
| | | | BW | GA | BW & GA | Sex | <22 weeks | ≥ 45 weeks | > 6,500g | Implausible BW | | BW | GA | BW & GA | Sex | < 22 weeks | ≥45 weeks | > 6,500 g | Implausible BW | |
| USA | 2019 | 3,747,984 | 0.1 | 0.1 | 0.0 | 0.0 | 0.1 | 0.0 | 0.0 | 0.0 | 21,556 | 6.9 | 0.5 | 0.2 | 3.3 | 22.3 | 0.0 | 0.0 | 0.2 | 14,724 |

** Scotland data- Late fetal deaths (LFD) at 22 or 23 weeks were classified as stillbirths for this study. Birthweight is a mandatory data item for stillbirths but not for LFD (22/23 weeks), hence most LFDs will not have a birthweight recorded.*

### Table S1c. Assessment of plausibility of input dataset 138 country-years from 125.4 million nationwide birth records, 2000 to 2020

**Calculated Late gestation Stillbirth Rate (SBR):** the number of stillbirths at 28 or more weeks of gestation divided by the number of total births at 28 or more weeks of gestation per 1000 calculated from the national dataset.

**National reported late gestation SBR:** SBR 28 or more weeks reported by the country to the United Nations Inter-Agency Group for Stillbirth Estimation (UNIGME)[3]

Difference= Calculated late gestation SBR – Reported late gestation SBR

| Country | Year | Total birth | Late Stillbirth (≥$28^{+0}$ weeks) | Calculated late gestation Stillbirth rate in input dataset | National reported late gestation Stillbirth rate | Difference |
|---|---|---|---|---|---|---|
| Argentina | 2017 | 629,578 | 3,505 | 5.6 | 5.2 | 0.4 |
| Argentina | 2018 | 611,150 | 3,465 | 5.7 | 5.4 | 0.3 |
| Estonia | 2015 | 13,908 | 38 | 2.7 | 3.1 | -0.4 |
| Estonia | 2016 | 13,865 | 36 | 2.6 | 2.8 | -0.2 |
| Estonia | 2017 | 13,507 | 35 | 2.6 | 2.6 | 0.0 |
| Estonia | 2018 | 14,146 | 29 | 2.1 | 2.2 | -0.1 |
| Estonia | 2019 | 13,865 | 20 | 1.4 | 1.8 | -0.4 |
| Estonia | 2020 | 13,003 | 25 | 1.9 | 1.9 | 0.0 |
| Malaysia | 2012 | 111,452 | 623 | 5.6 | 4.3 | 1.3 |
| Malaysia | 2013 | 111,572 | 577 | 5.2 | 4.3 | 0.9 |
| Malaysia | 2014 | 127,715 | 634 | 5.0 | 4.3 | 0.7 |
| Malaysia | 2015 | 132,189 | 654 | 4.9 | 4.4 | 0.5 |
| Malaysia | 2016 | 125,419 | 687 | 5.5 | 5.2 | 0.3 |
| Mexico | 2008 | 1,843,920 | 11,811 | 6.4 | 4.5 | 1.9 |
| Mexico | 2009 | 1,921,592 | 11,364 | 5.9 | 4.5 | 1.4 |
| Mexico | 2010 | 1,938,845 | 11,094 | 5.7 | 4.2 | 1.5 |
| Mexico | 2011 | 2,032,051 | 10,986 | 5.4 | 4.3 | 1.1 |
| Mexico | 2012 | 2,074,523 | 10,413 | 5.0 | 4.2 | 0.8 |
| Mexico | 2013 | 2,062,420 | 9,811 | 4.8 | 4 | 0.8 |
| Mexico | 2014 | 2,049,502 | 9,649 | 4.7 | 4 | 0.7 |
| Mexico | 2015 | 2,025,434 | 9,524 | 4.7 | 4.1 | 0.6 |

| | | | | | | |
|---|---|---|---|---|---|---|
| Mexico | 2016 | 1,967,312 | 8,904 | 4.5 | 3.9 | 0.6 |
| Mexico | 2017 | 1,952,806 | 8,870 | 4.5 | 4 | 0.5 |
| Mexico | 2018 | 1,854,178 | 9,316 | 5.0 | 3.9 | 1.1 |
| Netherlands | 2010 | 176,108 | 507 | 2.9 | 2.9 | 0.0 |
| Netherlands | 2011 | 174,077 | 510 | 2.9 | 3 | -0.1 |
| Netherlands | 2012 | 171,229 | 458 | 2.7 | 2.7 | 0.0 |
| Netherlands | 2013 | 166,481 | 389 | 2.3 | 2.2 | 0.1 |
| Netherlands | 2014 | 170,152 | 385 | 2.3 | 2.4 | -0.1 |
| Netherlands | 2015 | 165,196 | 394 | 2.4 | 2.4 | 0.0 |
| Netherlands | 2016 | 168,082 | 360 | 2.1 | 2.2 | -0.1 |
| Netherlands | 2017 | 164,718 | 343 | 2.1 | 1.9 | 0.2 |
| Netherlands | 2018 | 159,944 | 364 | 2.3 | 2 | 0.3 |
| Netherlands | 2019 | 162,896 | 390 | 2.4 | 2.3 | 0.1 |
| Netherlands | 2020 | 163,386 | 409 | 2.5 | 2.2 | 0.3 |
| Qatar | 2016 | 21,948 | 41 | 1.9 | 3.8 | -1.9 |
| Qatar | 2017 | 23,917 | 113 | 4.7 | 4.3 | 0.4 |
| Qatar | 2018 | 23,541 | 108 | 4.6 | 3.5 | 1.1 |
| Qatar | 2019 | 24,787 | 95 | 3.8 | 1.7 | 2.1 |
| Sweden | 2008 | 105,697 | 231 | 2.2 | 3 | -0.8 |
| Sweden | 2009 | 107,125 | 247 | 2.3 | 3 | -0.7 |
| Sweden | 2010 | 112,492 | 219 | 1.9 | 2.8 | -0.9 |
| Sweden | 2011 | 109,013 | 253 | 2.3 | 3 | -0.7 |

| | | | | | | |
|---|---|---|---|---|---|---|
| Sweden | 2012 | 109,909 | 253 | 2.3 | 2.9 | -0.6 |
| Sweden | 2013 | 110,350 | 237 | 2.1 | 2.8 | -0.7 |
| Sweden | 2014 | 112,849 | 277 | 2.5 | 3 | -0.5 |
| Sweden | 2015 | 113,231 | 247 | 2.2 | 2.8 | -0.6 |
| Sweden | 2016 | 115,865 | 210 | 1.8 | 2.7 | -0.9 |
| Sweden | 2017 | 113,928 | 234 | 2.1 | 2.7 | -0.6 |
| Sweden | 2018 | 114,648 | 259 | 2.3 | 3 | -0.7 |
| Sweden | 2019 | 113,284 | 219 | 1.9 | 2.4 | -0.5 |
| USA | 2000 | 3,995,629 | 11,334 | 2.8 | 3.3 | -0.5 |
| USA | 2001 | 3,967,327 | 11,222 | 2.8 | 3.3 | -0.5 |
| USA | 2002 | 3,961,263 | 10,957 | 2.8 | 3.2 | -0.4 |
| USA | 2003 | 4,026,035 | 10,737 | 2.7 | 3.1 | -0.4 |
| USA | 2004 | 4,049,000 | 10,820 | 2.7 | 3.1 | -0.4 |
| USA | 2005 | 4,088,130 | 10,606 | 2.6 | 3 | -0.4 |
| USA | 2006 | 4,219,682 | 10,896 | 2.6 | 3 | -0.4 |
| USA | 2007 | 4,291,613 | 11,032 | 2.6 | 3 | -0.4 |
| USA | 2008 | 4,226,361 | 11,251 | 2.7 | 3.2 | -0.5 |
| USA | 2009 | 4,110,306 | 10,474 | 2.5 | 2.9 | -0.4 |
| USA | 2010 | 3,980,108 | 10,266 | 2.6 | 3 | -0.4 |
| USA | 2011 | 3,935,233 | 10,278 | 2.6 | 3 | -0.4 |
| USA | 2012 | 3,934,778 | 10,121 | 2.6 | 3 | -0.4 |
| USA | 2013 | 3,915,272 | 10,114 | 2.6 | 3 | -0.4 |

| | | | | | | |
|---|---|---|---|---|---|---|
| USA | 2014 | 3,974,719 | 10,394 | 2.6 | 2.8 | -0.2 |
| USA | 2015 | 3,965,801 | 10,538 | 2.7 | 2.8 | -0.1 |
| USA | 2016 | 3,932,646 | 10,633 | 2.7 | 2.9 | -0.2 |
| USA | 2017 | 3,843,698 | 10,214 | 2.7 | 2.8 | -0.1 |
| USA | 2018 | 3,781,474 | 9,972 | 2.6 | 2.8 | -0.2 |
| USA | 2019 | 3,736,901 | 9,632 | 2.6 | 2.7 | -0.1 |

Source: [4]

### S2. RECORD guidelines checklist

| | # | STROBE items | Location | RECORD items | Location in manuscript where items are reported |
|---|---|---|---|---|---|
| | 1 | (a) Indicate the study's design with a commonly used term in the title or the abstract (b) Provide in the abstract an informative and balanced summary of what was done and what was found | | **RECORD 1.1:** The type of data used should be specified in the title or abstract. When possible, the name of the databases used should be included. **RECORD 1.2**: If applicable, the geographic region and timeframe within which the study took place should be reported in the title or abstract. **RECORD 1.3**: If linkage between databases was conducted for the study, this should be clearly stated in the title or abstract. | Title: "Stillbirths: contribution of newborn types according to attained size-for-gestational age in 13 countries using 125.1 million nationwide birth records, 2000 to 2020 " |
| Background rationale | 2 | Explain the scientific background and rationale for the investigation being reported | | | Introduction (Paragraphs 1-6) |
| Objectives | 3 | State specific objectives, including any prespecified hypotheses | | | Introduction (Paragraph 5-6) |
| Study Design | 4 | Present key elements of study design early in the paper | | | Methods (Paragraph 1-4) |
| Setting | 5 | Describe the setting, locations, and relevant dates, including periods of recruitment, exposure, follow-up, and data collection | | | Methods (Paragraphs 1-2) |
| Participants | 6 | *(a) Cohort study* - Give the eligibility criteria, and the sources and methods of selection of participants. Describe methods of follow-up *Case-control study* - Give the eligibility criteria, and the sources and methods of case ascertainment and control selection. Give the rationale for the choice of cases and controls. *Cross-sectional study* - Give the eligibility criteria, and the sources and methods of selection of participants. *(b) Cohort study* - For matched studies, give matching criteria and number of exposed and unexposed *Case-control study* - For matched studies, give matching criteria and the number of controls per case | | **RECORD 6.1:** The methods of study population selection (such as codes or algorithms used to identify subjects) should be listed in detail. If this is not possible, an explanation should be provided. **RECORD 6.2:** Any validation studies of the codes or algorithms used to select the population should be referenced. If validation was conducted for this study and not published elsewhere, detailed methods and results should be provided. **RECORD 6.3:** If the study involved linkage of databases, consider use of a flow diagram or other graphical display to demonstrate the data linkage process, including the number of individuals with linked data at each stage. | Methods (Paragraphs 2-3) under the subheading Inclusion and Exclusion criteria<br><br>Figure 1a. Flowchart |

| | # | STROBE items | Location | RECORD items | Location in manuscript where items are reported |
|---|---|---|---|---|---|
| Variables | 7 | Clearly define all outcomes, exposures, predictors, potential confounders, and effect modifiers. Give diagnostic criteria, if applicable. | | **RECORD 7.1:** A complete list of codes and algorithms used to classify exposures, outcomes, confounders, and effect modifiers should be provided. If these cannot be reported, an explanation should be provided. | Methods (Paragraphs 4) |
| Data sources/ measurement | 8 | For each variable of interest, give sources of data and details of methods of assessment (measurement). Describe comparability of assessment methods if there is more than one group | | | Methods (Paragraphs 4) |
| Bias | 9 | Describe any efforts to address potential sources of bias | | | Quality assessment described in Methods (Paragraph 3 and Supplementary material Table S1a) |
| Study size | 10 | Explain how the study size was arrived at | | | |
| Quantitative variables | 11 | Explain how quantitative variables were handled in the analyses. If applicable, describe which groupings were chosen, and why | | | Methods (Paragraphs 4) |
| Statistical methods | 12 | (a) Describe all statistical methods, including those used to control for confounding. (b) Describe any methods used to examine subgroups and interactions. (c) Explain how missing data were addressed. (d) *Cohort study* - If applicable, explain how loss to follow-up was addressed *Case-control study* - If applicable, explain how matching of cases and controls was addressed. *Cross-sectional study* - If applicable, describe analytical methods taking account of sampling strategy (e) Describe any sensitivity analyses | | | Methods (Paragraph 5-8) |
| Data access and cleaning methods | | .. | | **RECORD 12.1:** Authors should describe the extent to which the investigators had access to the database population used to create the study population. **RECORD 12.2:** Authors should provide information on the data cleaning methods used in the study. | Methods (Paragraph x) |

| | # | STROBE items | Location | RECORD items | Location in manuscript where items are reported |
|---|---|---|---|---|---|
| Linkage | | .. | | **RECORD 12.3:** State whether the study included person-level, institutional-level, or other data linkage across two or more databases. The methods of linkage and methods of linkage quality evaluation should be provided. | Supplementary |
| Participants | 13 | (a) Report the numbers of individuals at each stage of the study (*e.g.*, numbers potentially eligible, examined for eligibility, confirmed eligible, included in the study, completing follow-up, and analysed)<br>(b) Give reasons for non-participation at each stage.<br>(c) Consider use of a flow diagram | | **RECORD 13.1:** Describe in detail the selection of the persons included in the study (*i.e.,* study population selection) including filtering based on data quality, data availability and linkage. The selection of included persons can be described in the text and/or by means of the study flow diagram. | Results (Paragraph 1 and Figure 1) |
| Descriptive data | 14 | (a) Give characteristics of study participants (*e.g.*, demographic, clinical, social) and information on exposures and potential confounders.<br>(b) Indicate the number of participants with missing data for each variable of interest<br>(c) *Cohort study* - summarise follow-up time (*e.g.*, average and total amount) | | | |
| Outcome data | 15 | *Cohort study* - Report numbers of outcome events or summary measures over time<br>*Case-control study* - Report numbers in each exposure category, or summary measures of exposure<br>*Cross-sectional study* - Report numbers of outcome events or summary measures | | | |
| Main results | 16 | (a) Give unadjusted estimates and, if applicable, confounder-adjusted estimates and their precision (e.g., 95% confidence interval). Make clear which confounders were adjusted for and why they were included. (b) Report category boundaries when continuous variables were categorized.<br>(c) If relevant, consider translating estimates of relative risk into absolute risk for a meaningful time period | | | |

| | # | STROBE items | Location | RECORD items | Location in manuscript where items are reported |
|---|---|---|---|---|---|
| Other analyses | 17 | Report other analyses done—e.g., analyses of subgroups and interactions, and sensitivity analyses | | | |
| Key results | 18 | Summarise key results with reference to study objectives | | | |
| Limitations | 19 | Discuss limitations of the study, taking into account sources of potential bias or imprecision. Discuss both direction and magnitude of any potential bias | | **RECORD 19.1:** Discuss the implications of using data that were not created or collected to answer the specific research question(s). Include discussion of misclassification bias, unmeasured confounding, missing data, and changing eligibility over time, as they pertain to the study being reported. | |
| Interpretation | 20 | Give a cautious overall interpretation of results considering objectives, limitations, multiplicity of analyses, results from similar studies, and other relevant evidence | | | |
| Generalisability | 21 | Discuss the generalisability (external validity) of the study results | | | |
| Funding | 22 | Give the source of funding and the role of the funders for the present study and, if applicable, for the original study on which the present article is based | | | The source of founding is included in the Abstract and the funding role is described at the end of the manuscript under the subheading Funding role |
| Accessibility of protocol, raw data, and programming code | | .. | | **RECORD 22.1:** Authors should provide information on how to access any supplemental information such as the study protocol, raw data, or programming code. | Under the subheading Availability of data and material |

Source: Bechamel et al (2015)[4]

### S3. Ethics approval or exemptions of Institutional Review Boards

| Country of origin for data | Institutional Review Board(s) or data access provider | Ref/Number | Date of approval |
|---|---|---|---|
| London School of Hygiene & Tropical Medicine (LSHTM) | LSHTM - Observational / Interventions Research Ethics Committee | 22858 | 17th May 2021 |
| Estonia | Ethics Committee of National Institute for Health Development | 770 | 09th August 2021 |
| Iran | Iran University of Medical Sciences, Tehran, Iran | IR.IUMS.REC.1400.758 | 21st November 2021 |
| Lebanon | Institutional Review Board, American University of Beirut | PED.KY.01 | 13th July 2021 |
| Malaysia | Medical Research & Ethics Committee, Ministry of Health Malaysia | KKM/NIHSEC/ P21-718 (4) | 5the May 2021 |
| Mexico | Centre of Investigation in Health Sciences, Anahuac University, Mexico | 202214 | 31st March 2022 |
| Qatar | Medical Research Centre, Hamad Medical Corporation, Doha-Qatar | MRC-01-21-277 | 25th April 2021 |
| Kentland and Wales | 1. National Information Governance Board<br>2. Confidentiality Advisory Group of the Health Research Authority<br>3. Health & Social Care Information Centre (HSCIC), Data Access Advisory Group | 1. ECC 5-05 (f)/2012<br>2. 15/CAG/0119<br>3. DARS-NIC-359651-H3R1P-v5.2. | 10th October 2012 and 1st May2015 |
| UK Scotland | Public Health Scotland | 20210218-Vulnerable Newborn Measurement | 30th March 2021 |
| Exemptions (*e.g., IRB approval not required for public or aggregate data, existing ethics approval in place, etc)* | | | |
| Argentina | | | |
| Denmark | | | |
| Netherlands | | | |
| Sweden | | | |
| Uruguay | | | |
| USA publicly available data from https://www.cdc.gov/nchs/data_access/Vitalstatsonline.htm | | | |

### S4 Additional results

Table S4a. Stillbirth rate and rate ratio by newborn types for all gestation ($\geq 22^{+0}$ weeks)

| Country | Prevalence (%) (95%CI) | | | | | | Rate per 1,000 total births (95%CI) | | | | | | Rate ratio (95%CI) | | | | | |
|---|---|---|---|---|---|---|---|---|---|---|---|---|---|---|---|---|---|---|
| | PT+SGA | PT+AGA | PT+LGA | T+SGA | T+AGA | T+LGA | PT+SGA | PT+AGA | PT+LGA | T+SGA | T+AGA | T+LGA | PT+SGA | PT+AGA | PT+LGA | T+SGA | T+AGA | T+LGA |
| Argentina | 16.5<br>(16.2, 16.7) | 43.7<br>(41.5, 46) | 13.1<br>(11.2, 15.3) | 6.0<br>(5.8, 6.2) | 16.3<br>(15.6, 17) | 4.4<br>(3.6, 5.4) | 119<br>(119, 119) | 44.9<br>(44.9, 44.9) | 53.6<br>(53.5, 53.6) | 9.6<br>(9.5, 9.7) | 1.8<br>(1.7, 1.8) | 1.9<br>(1.7, 2.1) | 66.3<br>(66.2, 66.3) | 25<br>(25, 25.1) | 29.8<br>(29.7, 30) | 5.3<br>(5.3, 5.4) | 1<br>(Ref) | 1<br>(0.8, 1.3) |
| Denmark | 16<br>(16, 16) | 38.8<br>(38.8, 38.8) | 5.4<br>(5.4, 5.4) | 6.6<br>(6.6, 6.6) | 26<br>(26, 26) | 7.2<br>(7.2, 7.2) | 103<br>(103, 103) | 24.9<br>(24.9, 24.9) | 17.6<br>(17.6, 17.6) | 7.6<br>(7.6, 7.6) | 1.4<br>(1.4, 1.4) | 0.9<br>(0.9, 0.9) | 73.7<br>(73.7, 73.7) | 17.8<br>(17.8, 17.8) | 12.6<br>(12.6, 12.6) | 5.5<br>(5.5, 5.5) | 1<br>(Ref) | 0.7<br>(0.7, 0.7) |
| England & Wales | 21.4<br>(19.9, 23.1) | 45.3<br>(41.3, 49.7) | 4.4<br>(3.4, 5.8) | 5.7<br>(4, 8.1) | 19.5<br>(17.4, 21.9) | 3.4<br>(2.4, 4.8) | 97.4<br>(97.3, 97.6) | 31.6<br>(31.6, 31.7) | 23.9<br>(23.7, 24.2) | 5.8<br>(5.4, 6.2) | 1.2<br>(1, 1.4) | 0.8<br>(0.4, 1.2) | 81.6<br>(81.4, 81.7) | 26.5<br>(26.3, 26.7) | 20<br>(19.7, 20.3) | 4.9<br>(4.6, 5.1) | 1<br>(Ref) | 0.7<br>(0.3, 1) |
| Estonia | 13.8<br>(9.7, 19.5) | 37.5<br>(19.1, 73.5) | 4.5<br>(1.8, 11.3) | 4.7<br>(1.7, 12.9) | 27.1<br>(9.9, 74.7) | - | 93.9<br>(93.1, 94.6) | 22.9<br>(21.8, 24) | 15.3<br>(14.2, 16.5) | 7.4<br>(6.7, 8.1) | 1.3<br>(0.5, 2) | - | 74.5<br>(73.2, 75.8) | 18.2<br>(16.6, 19.8) | 12.2<br>(10.4, 13.9) | 5.9<br>(4.8, 7) | 1<br>(Ref) | - |
| Iran | 17.5<br>(15.3, 20.1) | 49.5<br>(47.1, 52) | 14.8<br>(12.2, 18) | 4.1<br>(3.9, 4.3) | 11.6<br>(10.4, 12.9) | 2.4<br>(2, 2.8) | 149.2<br>(149, 149.4) | 57.2<br>(57, 57.5) | 80.4<br>(80.3, 80.6) | 6<br>(5.6, 6.4) | 1.3<br>(0.9, 1.6) | 1.6<br>(1.3, 2) | 118.4<br>(118.2, 118.5) | 45.4<br>(45.3, 45.5) | 63.8<br>(63.6, 64) | 4.8<br>(4.7, 4.9) | 1<br>(Ref) | 1.3<br>(1.2, 1.4) |
| Malaysia | 28.8<br>(27.2, 30.4) | 36.3<br>(32.3, 40.9) | 6.9<br>(5, 9.5) | 13.1<br>(10.7, 16.1) | 12.5<br>(10.4, 15) | 2.2<br>(1.4, 3.4) | 126.5<br>(126.2, 126.7) | 31.7<br>(31.5, 32) | 21.7<br>(21.3, 22.2) | 6.7<br>(6.3, 7.1) | 1.3<br>(1, 1.6) | 2.9<br>(2.4, 3.5) | 95.3<br>(95.1, 95.5) | 23.9<br>(23.7, 24.2) | 16.4<br>(16, 16.7) | 5.1<br>(4.8, 5.3) | 1<br>(Ref) | 2.2<br>(1.8, 2.6) |
| Mexico | 0.9<br>(0.7, 1.2) | 56.4<br>(44.4, 71.6) | 11.5<br>(5.5, 24) | 3.2<br>(2.5, 4) | 21.8<br>(17.7, 26.8) | 4.1<br>(0.9, 18.2) | 11.4<br>(11.2, 11.5) | 77.3<br>(77.2, 77.4) | 161.7<br>(161, 162.3) | 3.9<br>(3.5, 4.3) | 2.2<br>(1.9, 2.5) | 3.7<br>(2.3, 5.2) | 5.2<br>(4.9, 5.5) | 35.3<br>(35.1, 35.5) | 73.8<br>(73.2, 74.4) | 1.8<br>(1.5, 2.1) | 1<br>(Ref) | 1.7<br>(0, 3.4) |
| The Netherlands | 17.9<br>(14.9, 21.5) | 50.9<br>(44.7, 58) | 5.3<br>(4, 7.2) | 5.3<br>(3.8, 7.4) | 16.5<br>(12.7, 21.5) | 3.5<br>(2.3, 5.4) | 131<br>(130.8, 131.2) | 42.7<br>(42.4, 42.9) | 31<br>(30.6, 31.4) | 8.3<br>(8, 8.6) | 1.2<br>(0.9, 1.5) | 0.7<br>(0.3, 1.2) | 109.5<br>(109.1, 109.8) | 35.6<br>(35.2, 36.1) | 25.9<br>(25.3, 26.4) | 7<br>(6.5, 7.4) | 1<br>(Ref) | 0.6<br>(0.1, 1.1) |
| Qatar | 28.0<br>(22.8, 34.3) | 40.3<br>(33, 49.2) | 4.4<br>(1.3, 15.2) | 6.1<br>(2.6, 14.5) | 16.1<br>(9.2, 28.2) | 2.0<br>(0.2, 17.8) | 132.3<br>(131.4, 133.2) | 25.4<br>(24.3, 26.6) | 15.6<br>(14.9, 16.4) | 5.4<br>(3.6, 7.2) | 1.1<br>(0.3, 2) | 0.8<br>( -0.5, 2.1) | 117.4<br>(116.6, 118.1) | 22.6<br>(21.9, 23.2) | 13.9<br>(12.5, 15.2) | 4.8<br>(3.4, 6.2) | 1<br>(Ref) | 0.7<br>( -1.1, 2.5) |
| Scotland | 16.2<br>(12.8, 20.5) | 32.0<br>(26.1, 39.2) | 4.5<br>(2.2, 9.1) | 10.6<br>(6.3, 18) | 29.5<br>(22.9, 37.9) | 5.9<br>(3.3, 10.5) | 94.8<br>(94.3, 95.2) | 23.1<br>(22.7, 23.6) | 22.2<br>(21.3, 23) | 10.8<br>(10.3, 11.3) | 2<br>(1.6, 2.3) | 1.3<br>(0.5, 2.1) | 48.5<br>(48.1, 48.9) | 11.8<br>(11.5, 12.2) | 11.4<br>(10.5, 12.2) | 5.5<br>(5.1, 6) | 1<br>(Ref) | 0.7<br>( -0.1, 1.4) |
| Sweden | 16.2<br>(11.7, 22.4) | 31.1<br>(27.4, 35.4) | 6.8<br>(4.7, 9.9) | 9.2<br>(7, 12) | 28.9<br>(25.4, 32.9) | 7.1<br>(4.7, 10.7) | 85.5<br>(85.1, 85.9) | 19.9<br>(19.7, 20.1) | 35.8<br>(35.5, 36.2) | 8<br>(7.6, 8.3) | 1.3<br>(1.1, 1.4) | 0.9<br>(0.5, 1.3) | 67.9<br>(67.5, 68.3) | 15.8<br>(15.6, 16.0) | 28.4<br>(28.1, 28.8) | 6.3<br>(6, 6.7) | 1<br>(1, 1) | 0.7<br>(0.3, 1.2) |
| USA | 23.7<br>(22.6, 24.8) | 44.2<br>(42.4, 46.1) | 8.6<br>(7.8, 9.4) | 5.7<br>(4.8, 6.8) | 13.8<br>(12.9, 14.9) | 3.9<br>(3.5, 4.4) | 118.4<br>(118.4, 118.5) | 23.2<br>(23.1, 23.3) | 22<br>(21.8, 22.3) | 5.8<br>(5.6, 5.9) | 0.8<br>(0.7, 1) | 0.9<br>(0.8, 1) | 140.9<br>(140.8, 141) | 27.6<br>(27.5, 27.7) | 26.2<br>(25.9, 26.5) | 6.9<br>(6.8, 7) | 1<br>(Ref) | 1.1<br>(0.9, 1.2) |

### Table S4b. Stillbirth rate by newborn types for late gestation ($\geq 28^{+0}$ weeks)

| Country | Prevalence (%) (95%CI) | | | | | | Rate per 1,000 total births (95%CI) | | | | | | Rate ratio (95%CI) | | | | | |
|---|---|---|---|---|---|---|---|---|---|---|---|---|---|---|---|---|---|---|
| | PT+SGA | PT+AGA | PT+LGA | T+SGA | T+AGA | T+LGA | PT+SGA | PT+AGA | PT+LGA | T+SGA | T+AGA | T+LGA | PT+SGA | PT+AGA | PT+LGA | T+SGA | T+AGA | T+LGA |
| Argentina | 17.1 (17, 17.1) | 37 (35.1, 38.9) | 10.3 (9.1, 11.8) | 8 (7.9, 8.2) | 21.7 (21.2, 22.2) | 5.9 (4.8, 7.3) | 98 (98, 98.1) | 30.2 (30.1, 30.2) | 33.5 (33.5, 33.6) | 9.6 (9.5, 9.7) | 1.8 (1.7, 1.8) | 1.9 (1.7, 2.1) | 54.6 (54.6, 54.6) | 16.8 ( 16.8 , 16.8 ) | 18.7 (18.6, 18.8) | 5.3 (5.3, 5.4) | 1 (Ref) | 1 (0.8, 1.3) |
| Denmark | 12.6 (12.6, 12.6) | 27.7 (27.7, 27.7) | 4.2 (4.2, 4.2) | 9.2 (9.2, 9.2) | 36.3 (36.3, 36.3) | 10.1 (10.1, 10.1) | 61.9 (61.9, 61.9) | 13.5 (13.5, 13.5) | 10.3 (10.3, 10.3) | 7.6 (7.6, 7.6) | 1.4 (1.4, 1.4) | 0.9 (0.9, 0.9) | 44.3 (44.3, 44.3) | 9.7 ( NA , NA ) | 7.3 (7.3, 7.3) | 5.5 (5.5, 5.5) | 1 (Ref) | 0.7 (0.7, 0.7) |
| England & Wales | 18.1 (16.1, 20.4) | 34.2 (30.9, 37.8) | 3.6 (2.8, 4.7) | 8.7 (6.2, 12) | 29.9 (27.7, 32.2) | 5.2 (3.8, 7.3) | 61.3 (61.2, 61.4) | 17 (16.9, 17.1) | 13.7 (13.4, 14) | 5.8 (5.4, 6.2) | 1.2 (1, 1.4) | 0.8 (0.4, 1.2) | 51.3 (51.1, 51.5) | 14.2 ( 14 , 14.4 ) | 11.4 (11.2, 11.7) | 4.9 (4.6, 5.1) | 1 (Ref) | 0.7 (0.3, 1) |
| Estonia | 20.2 (6.3, 65.2) | - | - | 6.4 (2.4, 17.4) | 37 (15.9, 86) | - | 114.1 (112.7, 115.5) | - | - | 7.4 (6.7, 8.1) | 1.3 (0.5, 2) | - | 90.6 (88.7, 92.4) | - | - | 5.9 (4.8, 7) | 1 (Ref) | - |
| Iran | 18.4 (16.5, 20.6) | 35.2 (32.9, 37.6) | 10.8 (9.2, 12.7) | 8.1 (7.7, 8.4) | 22.7 (20.9, 24.7) | 4.7 (4.1, 5.4) | 96.7 (96.5, 96.9) | 23.3 (23, 23.6) | 34.6 (34.4, 34.9) | 6 (5.6, 6.4) | 1.3 (0.9, 1.6) | 1.6 (1.3, 2) | 76.7 (76.6, 76.8) | 18.4 ( 18.4 , 18.5 ) | 27.5 (27.4, 27.6) | 4.8 (4.7, 4.9) | 1 (Ref) | 1.3 (1.2, 1.4) |
| Malaysia | 28.9 (26.4, 31.6) | 29.2 (25.1, 34) | 5.4 (3.7, 7.9) | 17 (14, 20.7) | 16.2 (13.8, 19.1) | 2.8 (1.9, 4.3) | 101.8 (101.5, 102) | 20.5 (20.3, 20.7) | 14.4 (13.9, 14.9) | 6.7 (6.3, 7.1) | 1.3 (1, 1.6) | 2.9 (2.4, 3.5) | 76.7 (76.5, 76.9) | 15.4 ( 15.2 , 15.7 ) | 10.8 (10.5, 11.2) | 5.1 (4.8, 5.3) | 1 (Ref) | 2.2 (1.8, 2.6) |
| Mexico | 1 (0.9, 1.2) | 48.2 (42.3, 55.1) | 4.3 (2.2, 8.2) | 4.9 (4.1, 5.8) | 33.4 (27.5, 40.6) | 6.3 (1.4, 28.1) | 8.5 (8.2, 8.8) | 46 (45.8, 46.2) | 46.1 (45.4, 46.8) | 3.9 (3.5, 4.3) | 2.2 (1.9, 2.5) | 3.7 (2.3, 5.2) | 3.9 (3.7, 4.1) | 21 ( 20.8 , 21.1 ) | 21 (20.2, 21.8) | 1.8 (1.5, 2.1) | 1 (Ref) | 1.7 (0, 3.4) |
| Netherlands | 13.8 (10.7, 18) | 29.9 (26, 34.5) | 4.7 (2.6, 8.6) | 10.6 (8, 14) | 33.1 (27.8, 39.4) | 7.1 (4.7, 10.7) | 58 (57.7, 58.3) | 13.6 (13.4, 13.9) | 14.8 (14.2, 15.4) | 8.3 (8, 8.6) | 1.2 (0.9, 1.5) | 0.7 (0.3, 1.2) | 48.5 (48.2, 48.7) | 11.4 ( 11.1 , 11.7 ) | 12.4 (11.6, 13.1) | 7 (6.5, 7.4) | 1 (Ref) | 0.6 (0.1, 1.1) |
| Qatar | 25.2 (19.3, 32.9) | 28.6 (18.8, 43.4) | 6.3 (1.2, 32.2) | 8.7 (3.5, 21.6) | 22.9 (12.7, 41.4) | 2.8 (0.3, 24.2) | 91 (90.2, 91.8) | 13.6 (12.4, 14.8) | 16.8 (15.1, 18.5) | 5.4 (3.6, 7.2) | 1.1 (0.3, 2) | 0.8 ( -0.5, 2.1) | 80.7 (79.8, 81.6) | 12 ( 11.3 , 12.8 ) | 14.9 (13, 16.8) | 4.8 (3.4, 6.2) | 1 (Ref) | 0.7 ( -1.1, 2.5) |
| Scotland | 14 (10.3, 19) | 28 (22.1, 35.4) | 4.1 (2, 8.4) | 12.1 (7.3, 20.2) | 33.6 (26.7, 42.4) | 6.7 (3.7, 12.2) | 75.9 (75.3, 76.4) | 18.7 (18.2, 19.1) | 18.7 (17.8, 19.6) | 10.8 (10.3, 11.3) | 2 (1.6, 2.3) | 1.3 (0.5, 2.1) | 38.8 (38.4, 39.3) | 9.6 ( 9.2 , 10 ) | 9.6 (8.7, 10.4) | 5.5 (5.1, 6) | 1 (Ref) | 0.7 ( -0.1, 1.4) |
| Sweden | 5.3 (1.5, 18.2) | 28.6 (22.9, 35.6) | - | 12.6 (9.8, 16.1) | 39.8 (33, 47.9) | 9.8 (6.4, 14.8) | 22.9 (21.7, 24.2) | 14 (13.7, 14.4) | - | 8 (7.6, 8.3) | 1.3 (1.1, 1.4) | 0.9 (0.5, 1.3) | 18.2 (16.9, 19.5) | 11.1 ( 10.8 , 11.5 ) | - | 6.3 (6, 6.7) | 1 (Ref) | 0.7 (0.3, 1.2) |
| USA | 19.8 (18.4, 21.3) | 36.1 (32.7, 39.8) | 7.3 (6.7, 8) | 8.9 (7.4, 10.7) | 21.6 (20.3, 23.1) | 6.1 (5.5, 6.9) | 72.2 (72, 72.3) | 13 (12.9, 13.1) | 12.8 (12.4, 13.1) | 5.8 (5.6, 5.9) | 0.8 (0.7, 1) | 0.9 (0.8, 1) | 85.8 (85.7, 85.9) | 15.5 ( 15.3 , 15.6 ) | 15.2 (14.8, 15.6) | 6.9 (6.8, 7) | 1 (Ref) | 1.1 (0.9, 1.2) |

### S5. Summary of metadata

| Country | Units for recording | Data source for live and stillbirths | Reporting criteria for births | | | | | |
|---|---|---|---|---|---|---|---|---|
| | Birthweight (e.g.,gram/ oz' lb)/ Gestational age (e.g., weeks, days) | And details of linkage where relevant | Livebirths: Exclusions criteria based on BW | Livebirths: Exclusion criteria based on GA | Stillbirths: Exclusions criteria based on BW | Stillbirths: Exclusion criteria based on GA | Legal requirement for stillbirth reporting | Are births following induced Termination of Pregnancy included in the data source? |
| Argentina | Grams<br>Completed weeks<br>Completed weeks | | There are no birthweight limits if the baby was born alive | There are no gestational age limits if the baby was born alive | | | | They are included among the stillbirths |
| Denmark | Grams<br>Completed weeks | Information on livebirths and stillbirths were extracted from the Danish Medical Birth Registry. | None | | | <22 weeks[5] | | No |
| England & Wales | Grams<br>Weeks + days | Data linkage with birth notifications, and birth and stillbirth registrations | None | | | 22 weeks | | No |
| Estonia | Grams<br>Days | Estonian Medical Birth Register | None | | | <22 weeks. Abortions can be made up to 21 weeks and 6 days only in case of medical indication, all stillbirths (i.e. dead before expulsion or extraction ) are | | No |

| Country | Units for recording | Data source for live and stillbirths | Reporting criteria for births | | | | | |
|---|---|---|---|---|---|---|---|---|
| | Birthweight (e.g.,gram/ oz' lb)/ Gestational age (e.g., weeks, days) | And details of linkage where relevant | Livebirths: Exclusions criteria based on BW | Livebirths: Exclusion criteria based on GA | Stillbirths: Exclusions criteria based on BW | Stillbirths: Exclusion criteria based on GA | Legal requirement for stillbirth reporting | Are births following induced Termination of Pregnancy included in the data source? |
| | | | | | | included from 22 weeks onwards<br>. | | |
| Iran | | | | | | | | |
| Lebanon | Grams<br>Weeks + days | None | None | | | <22 weeks | | No |
| Malaysia | Grams<br>Completed weeks | None | None | | | None | | No |
| Mexico | Grams<br>Completed weeks | Livebirths and deaths records were linked using the variables sex, date of birth, place of residence and place of occurrence using CIDACS-RL software | None | | | None | | No |
| Netherlands | Grams<br>Completed weeks | | Gestational age <22 weeks; if ga. missing, birthweight<500 g | | | Gestational age <22 weeks; if ga. missing, birthweight<500 g | | Both livebirths and stillbirths up to 24 weeks gestation may include termination of pregnancy |
| Qatar | Kilograms and grams<br>Weeks and days | | None | | | None | | Yes, all births included |
| Scotland | Grams<br>Completed weeks | Live, stillbirth (>=24wks) and late fetal death (22/23 wks.) records were derived | None | | | None | Stillbirths (>=24 weeks) must be registered with | Termination of pregnancy records are excluded. |

| Country | Units for recording | Data source for live and stillbirths | Reporting criteria for births | | | | | |
|---|---|---|---|---|---|---|---|---|
| | Birthweight (e.g.,gram/ oz’ lb)/ Gestational age (e.g., weeks, days) | And details of linkage where relevant | Livebirths: Exclusions criteria based on BW | Livebirths: Exclusion criteria based on GA | Stillbirths: Exclusions criteria based on BW | Stillbirths: Exclusion criteria based on GA | Legal requirement for stillbirth reporting | Are births following induced Termination of Pregnancy included in the data source? |
| | | from SMR02 (hospital IP/DC hospital return for obstetric data). NRS stillbirths and NRS infant deaths (statutory national birth and death registers) were then matched to the SMR02 file using the Mother UPI number (unique ID), date of delivery and birth order. SMR02 is a file based on babies (live and stillbirths) was matched with NHS Lothian file and NRS infant deaths | | | | | National Records of Scotland. Late fetal deaths (22/23 weeks) are not registered. | |
| Sweden | Grams<br>Days | National databases that are linked together using the person-unique national registration numbers of children | None | | | <23 weeks | In Sweden information on stillbirths is available from 28 weeks onward from 1997 to July 1, 2008, and thereafter from 22 gestational weeks | Included after 22 weeks |
| Uruguay | Grams<br>Completed weeks | | None | | | None | | No |
| United States of America | Grams (and lb converted to | Data on live births and fetal deaths are available from | None | <20 weeks[5] | | | | The NCHS recommendation for fetal death definition is |

| Country | Units for recording | Data source for live and stillbirths | Reporting criteria for births | | | | | |
|---|---|---|---|---|---|---|---|---|
| | Birthweight (e.g.,gram/ oz' lb)/ Gestational age (e.g., weeks, days) | And details of linkage where relevant | Livebirths: Exclusions criteria based on BW | Livebirths: Exclusion criteria based on GA | Stillbirths: Exclusions criteria based on BW | Stillbirths: Exclusion criteria based on GA | Legal requirement for stillbirth reporting | Are births following induced Termination of Pregnancy included in the data source? |
| | grams)<br>Completed weeks | the NCHS – Vital Statistics online. It includes data from live birth certificates and fetal death certificates. | | | | | | to exclude TOPs. Some states do exclude TOPs regardless of gestational age, while some states include TOPs |
